# Integrative proteomic analyses across common cardiac diseases yield new mechanistic insights and enhanced prediction

**DOI:** 10.1101/2023.12.19.23300218

**Authors:** Art Schuermans, Ashley B. Pournamdari, Jiwoo Lee, Rohan Bhukar, Shriienidhie Ganesh, Nicholas Darosa, Aeron M. Small, Zhi Yu, Whitney Hornsby, Satoshi Koyama, James L. Januzzi, Michael C. Honigberg, Pradeep Natarajan

## Abstract

Cardiac diseases represent common highly morbid conditions for which underlying molecular mechanisms remain incompletely understood. Here, we leveraged 1,459 protein measurements in 44,313 UK Biobank participants to characterize the circulating proteome associated with incident coronary artery disease, heart failure, atrial fibrillation, and aortic stenosis. Multivariable-adjusted Cox regression identified 820 protein-disease associations—including 441 proteins—at Bonferroni-adjusted *P*<8.6×10^−6^. *Cis*-Mendelian randomization suggested causal roles that aligned with epidemiological findings for 6% of proteins identified in primary analyses, prioritizing novel therapeutic targets for different cardiac diseases (e.g., interleukin-4 receptor for heart failure and spondin-1 for atrial fibrillation). Interaction analyses identified seven protein-disease associations that differed Bonferroni-significantly by sex. Models incorporating proteomic data (vs. clinical risk factors alone) improved prediction for coronary artery disease, heart failure, and atrial fibrillation. These results lay a foundation for future investigations to uncover novel disease mechanisms and assess the clinical utility of protein-based prevention strategies for cardiac diseases.

## Main

Cardiac diseases represent the leading global cause of morbidity and mortality^1^, with coronary artery disease, heart failure, atrial fibrillation, and aortic stenosis collectively accounting for more than 90% of cardiac deaths^1,2^. The prevention of these diseases often relies on risk assessment and early pharmacotherapy, yet most high-risk individuals remain undetected until they experience their first clinical event^3,4^. In addition, even under optimal treatment conditions, there remains significant residual risk that is incompletely captured by traditional risk factors^5,6^. The development of effective screening tools and discovery of novel therapeutic targets could significantly enhance early detection and improve treatment outcomes for different cardiac diseases.

Large-scale genetic analyses have identified thousands of sequence variants associated with cardiac diseases, enabling insights into disease etiology and facilitating the development of risk prediction tools^7–11^. However, for many genetic associations, molecular effectors remain incompletely understood. For instance, recent genome-wide association studies of coronary artery disease suggest that only ∼50% of identified genetic loci confer risk through known risk factors such as lipids, obesity, and blood pressure^7,8^. Molecular pathways through which the remaining loci (with signals typically residing in non-coding DNA sequence) exert their effects remain less clear. However, they ultimately may represent ideal potential targets to reduce residual cardiovascular disease risk.

The circulating proteome is a dynamic network that encompasses all proteins in the bloodstream, reflecting both genetic background and external factors such as environmental exposures and lifestyle alterations. Smaller studies have demonstrated that sparse protein-based risk scores can improve the prediction of recurrent events in individuals with established atherosclerotic cardiovascular disease^12,13^. Furthermore, targeted analyses of specific biomarkers suggest that integrating proteomic and genetic data can nominate causal protein-disease associations and reveal actionable drug targets in the bloodstream^14,15^. Whether population-scale, agnostic analyses of the circulating proteome can provide novel insights into disease biology and improve the prediction of first clinical events across a range of cardiac disease subtypes remains unclear.

In the present study, we perform a proteomic analysis of cardiac diseases in the UK Biobank Pharma Proteomics Project (UKB-PPP)^16^, a population-based cohort including 54,306 individuals with longitudinal follow-up (**Fig. 1**). Using multivariable-adjusted time-to-event models, we test the associations of 1,459 circulating proteins with incident coronary artery disease, heart failure, atrial fibrillation, and aortic stenosis. We leverage Mendelian randomization (MR) to infer causal roles among the identified proteins to prioritize novel therapeutic targets. Finally, we present sex-specific protein-disease associations and evaluate whether protein-based scores can improve disease prediction beyond the use of traditional risk factors.

**Fig. 1.**
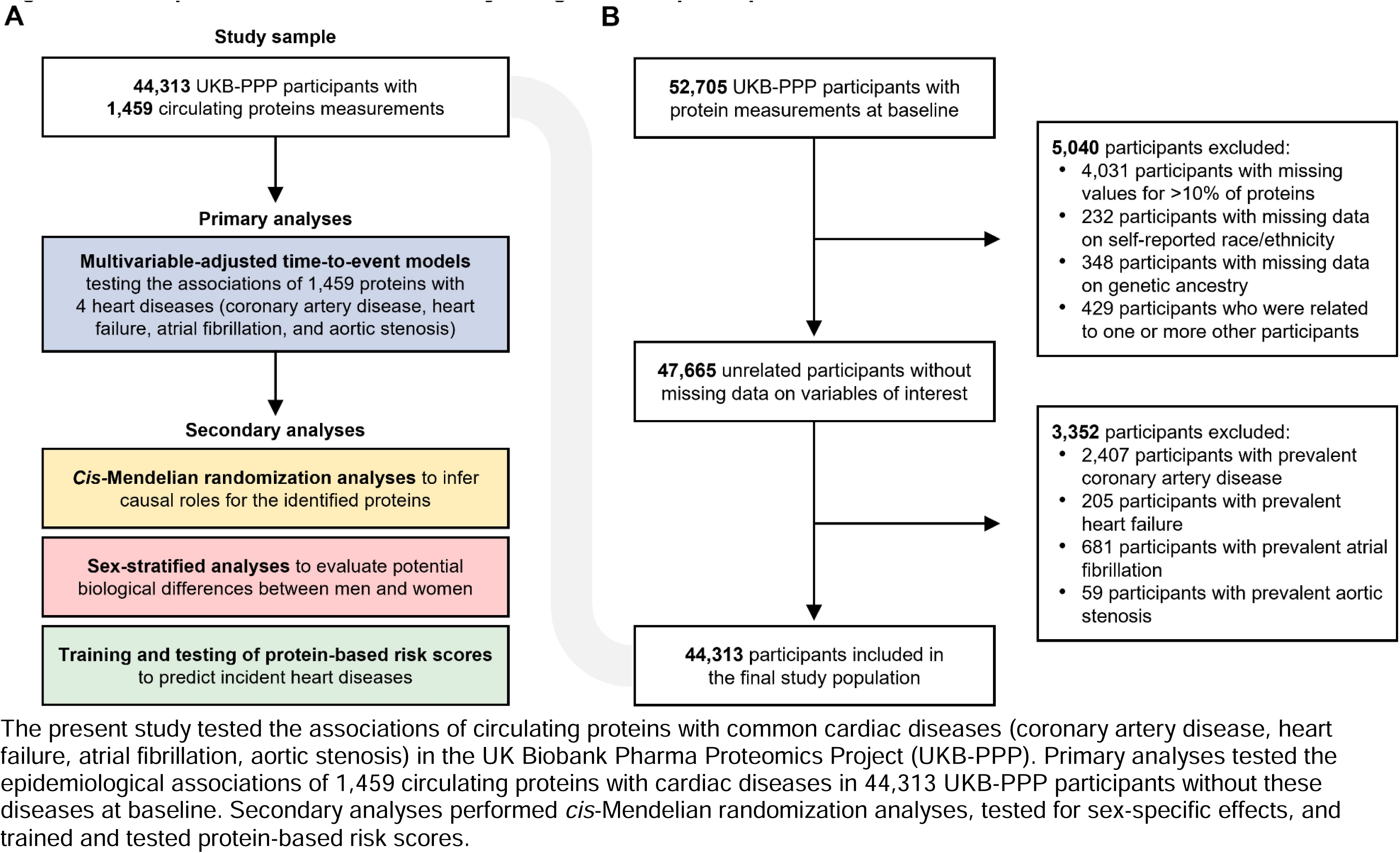
Visual representation of the (A) study design and (B) participant inclusion and exclusion criteria.

## Results

### Study population

The present study included 44,313 unrelated UKB-PPP participants without a history of coronary artery disease, heart failure, atrial fibrillation, or aortic stenosis at enrolment. Participant characteristics are summarized in **Table 1**. Baseline blood samples underwent proteomic profiling using the *Olink Explore 1536* platform, providing data on 1,459 unique proteins (**Supplementary Table 1**) available for >90% of UKB-PPP participants (**Supplementary Table 2**).

**Table 1.** Baseline characteristics of UK Biobank Pharma Proteomics Project (UKB-PPP) participants included in the present study (*N*=44,313).

|  | UKB-PPP participants (N=44,313) |
| --- | --- |
| Age at blood draw, y | 56.4 ± 8.2 |
| Female, *n* | 24,701 (55.7%) |
| Race/ethnicity, *n* | – |
| Asian | 942 (2.1%) |
| Black | 1,051 (2.4%) |
| White | 41,481 (93.6%) |
| Mixed | 300 (0.7%) |
| Other | 539 (1.2%) |
| Smoking status, *n* | – |
| Never | 24,709 (55.8%) |
| Previous | 14,932 (33.7%) |
| Current | 4,672 (10.5%) |
| Body mass index, kg/m<sup>2</sup> | 27.3 ± 4.7 |
| Blood pressure, mmHg | – |
| Systolic blood pressure | 139.5 ± 19.6 |
| Diastolic blood pressure | 82.3 ± 10.6 |
| Blood biochemistry, mg/dL | – |
| Total cholesterol | 221.4 ± 43.8 |
| LDL cholesterol | 138.3 ± 33.4 |
| HDL cholesterol | 56.4 ± 14.7 |
| Triglycerides | 129.8 (92.0 to 188.1) |
| Creatinine | 0.82 ± 0.19 |
| Type 2 diabetes mellitus, *n* | 1,218 (2.7%) |
| Medication use, *n* | – |
| Cholesterol-lowering medication use | 6,544 (14.8%) |
| Antihypertensive medication use | 6,214 (14.0%) |
| Townsend deprivation index | -2.08 (-3.63 to 0.70) |

Continuous variables are summarized as mean ± standard deviation or median (interquartile range), as appropriate. Categorical variables are summarized as *n* (%).

The majority of participants were female (*n*=24,701 [55.7%]) and self-reported as white (*n*=41,481 [93.6%]), with a mean (standard deviation [SD]) age of 56.4 (8.2) years. A total of 4,610 participants (10.4%) experienced at least one cardiac event over a median (interquartile range, IQR) follow-up of 11.1 (10.4-11.8) years. Coronary artery disease had the highest cumulative incidence (6.2% [*n*=2,729/44,313]), followed by atrial fibrillation (4.8% [*n*=2,107/44,313]), heart failure (2.3% [*n*=1,014/44,313]), and aortic stenosis (0.7% [*n*=326/44,313]) (**Supplementary** Fig. 1).

### Associations of circulating proteins with incident heart diseases

Primary analyses tested the associations of 1,459 circulating proteins with each of the incident heart diseases (coronary artery disease, heart failure, atrial fibrillation, and aortic stenosis) using multivariable-adjusted Cox regression models. These analyses identified 820 protein-disease associations—reflecting 441 unique proteins—at Bonferroni-corrected *P*<0.05/5,836 (**Fig. 2, Supplementary Table 3**). Heart failure had the highest number of proteomic associations (*n*=384), followed by coronary artery disease (*n*=259), atrial fibrillation (*n*=156), and aortic stenosis (*n*=21). Among proteins with one or more significant associations, 261 (59.2%) were shared across multiple outcomes and 15 (3.4%) were shared across all four outcomes (**Supplementary** Fig. 2).

**Fig. 2.**
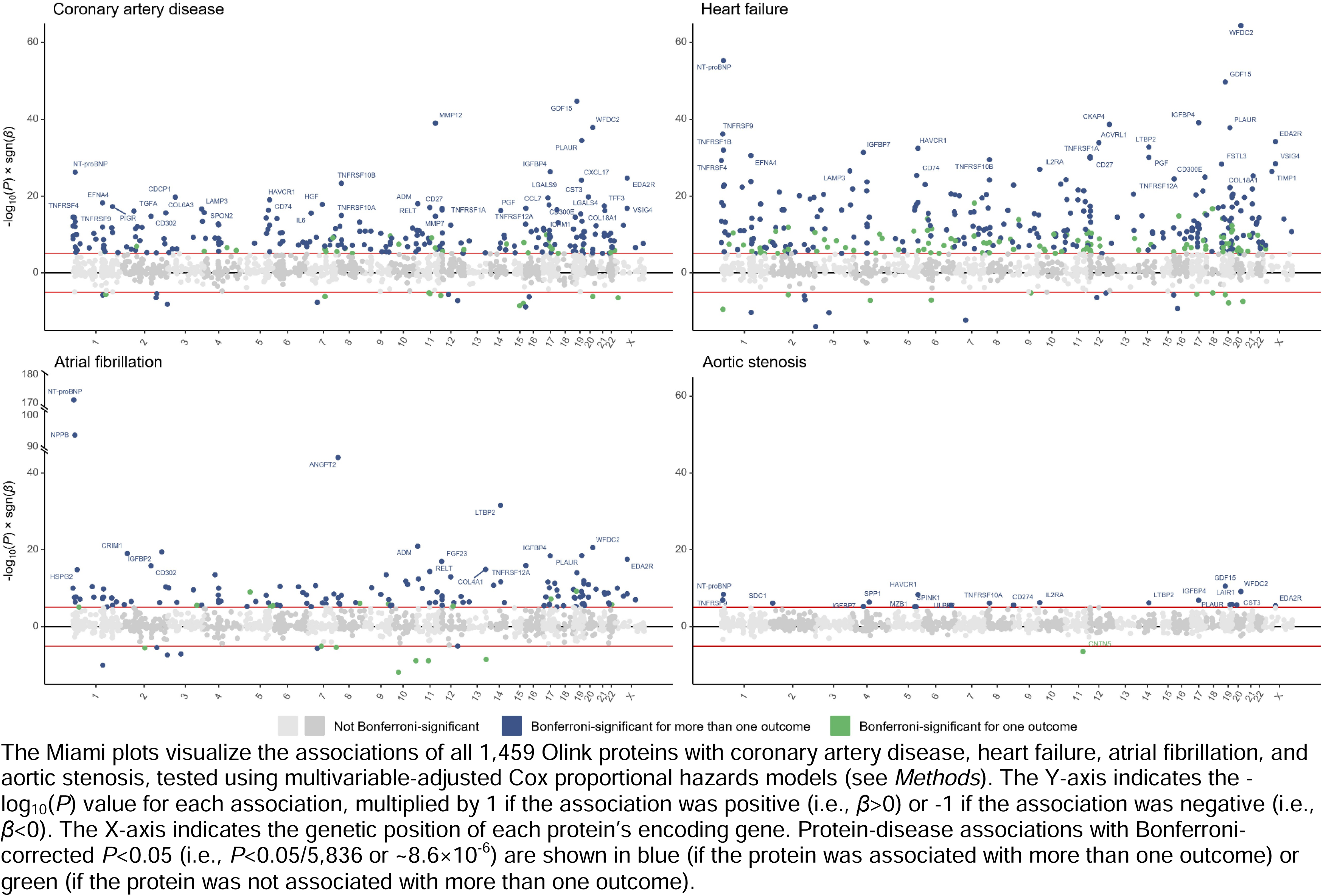
Associations of circulating protein levels with incident coronary artery disease, heart failure, atrial fibrillation, and aortic stenosis.

The strongest protein-disease associations (by *P*-value) were observed for atrial fibrillation, with N-terminal pro-B type natriuretic peptide (NT-proBNP) and B-type natriuretic peptide (NPPB, also known as BNP) yielding hazard ratios (HRs) of 1.74 (95% confidence interval, CI: 1.67-1.81; *P*=8.7×10^−173^) and 1.62 (95%CI: 1.54-1.69; *P*=5.6×10^−95^), respectively, for each SD increase in circulating protein levels. NT-proBNP was also the second strongest for association with heart failure, with an HR of 1.57 (95%CI: 1.48-1.66; *P*=5.4×10^−56^) per SD. The biomarker most strongly associated with heart failure was WAP four-disulfide core domain protein 2 (WFDC2), a fibroblast-derived mediator of fibrosis also known as human epididymis protein 4 (HE4)^17^, with an HR of 1.62 (95%CI: 1.54-1.72; *P*=4.1×10^−65^) per SD. The proteins most strongly associated with incident coronary artery disease were growth differentiation factor 15 (GDF15; HR, 1.31 [95%CI: 1.26-1.36] per SD; *P*=2.0×10^−45^) and matrix metalloproteinase-12 (MMP12; HR, 1.29 [95%CI: 1.24-1.34] per SD; *P*=1.1×10^−39^); those most strongly associated with aortic stenosis were GDF15 (HR, 1.44 [95%CI: 1.29-1.60] per SD; *P*=2.7×10^−11^) and WFDC2 (HR, 1.40 [95%CI: 1.26-1.55] per SD; *P*=7.3×10^−10^).

To gain insights into biological pathways associated with the identified proteins, we carried the 820 observed protein-disease associations forward for pathway enrichment analysis using the *Gene Ontology* resource^18^ via *Enrichr*^19^. The highest-scoring pathways for coronary artery disease and heart failure included inflammatory and immune-related processes involving leukocyte/lymphocyte chemotaxis and cellular response to cytokines (**Supplementary** Fig. 3, **Supplementary Table 4**). Participants with coronary artery disease or heart failure during follow-up were also enriched for apoptosis-related proteins such as those from the tumor necrosis factor (TNF) receptor family. Proteins associated with aortic stenosis demonstrated enrichment for peptidase inhibitor activity, consistent with recent work suggesting an important role for certain peptidases in the progression of calcific aortic stenosis^20^. Significantly enriched cellular components included the endoplasmatic reticulum lumen (for all cardiac outcomes), collagen-containing extracellular matrix (for coronary artery disease, heart failure, and atrial fibrillation), and intracellular organelle lumen (for atrial fibrillation and aortic stenosis).

### Cis-Mendelian randomization analyses

Next, we performed *cis*-MR analyses to infer causal effects of the identified proteins on coronary artery disease, heart failure, atrial fibrillation, and aortic stenosis. Of 441 unique Bonferroni-significant proteins in primary analyses, 430 (97.5%; corresponding to 802 protein-disease associations) had at least one valid *cis*-pQTL (±200 kilobases, *P*<5×10^−6^, *R*²<0.1) (**Supplementary Table 5**). *F*-statistics were >10 for all proteins other than myoglobin, which was excluded from *cis*-MR analyses to minimize the risk of weak instrument bias. Median (IQR) *F*-statistics and *R*² estimates (representing phenotypic variance explained by genetic instruments) were 1515 (454-4050) and 3.2% (1.0-7.9%), respectively (**Supplementary Table 6**). Consistent with the use of *cis*-pQTLs^21^, Steiger filtering did not identify any variants explaining more variance in the outcome than the exposure (**Supplementary Table 7**).

Of 801 protein-disease associations examined in *cis*-MR analyses, 225 (28.1%; representing 172/429 [40.1%] proteins) showed suggestive evidence of causality with *P*<0.05 (**Fig. 3**, **Supplementary Table 8**). Because it is routinely recommended to evaluate *cis*-MR findings across *P*-value and *R*² thresholds^15^, we performed multiple sensitivity analyses (using genetic variants at *P*<5×10^−4^/*<*5×10^−6^/<5×10^−8^ and *R*²<0.001/<0.01/<0.1/<0.2) to evaluate the robustness of the observed genetic associations (**Supplementary Table 9**). We further performed MR-Egger (**Supplementary Table 9**), one-sample MR (**Supplementary Table 10**), and multivariable-adjusted MR adjusting for proteins with shared pQTLs (**Supplementary Tables 11-12**). A total of 97/225 (43.1%) genetic protein-disease associations were robust across all sensitivity analyses. Genetic and observational analyses showed directional consistency for 48/97 (49.5%) robust genetic associations, corresponding to 6.0% of all protein-disease pairs tested in *cis*-MR analyses. These protein-disease associations all had positive effect estimates, implying that increased protein concentrations may promote cardiac disease risk and lowering would reduce risk.

**Fig. 3.**
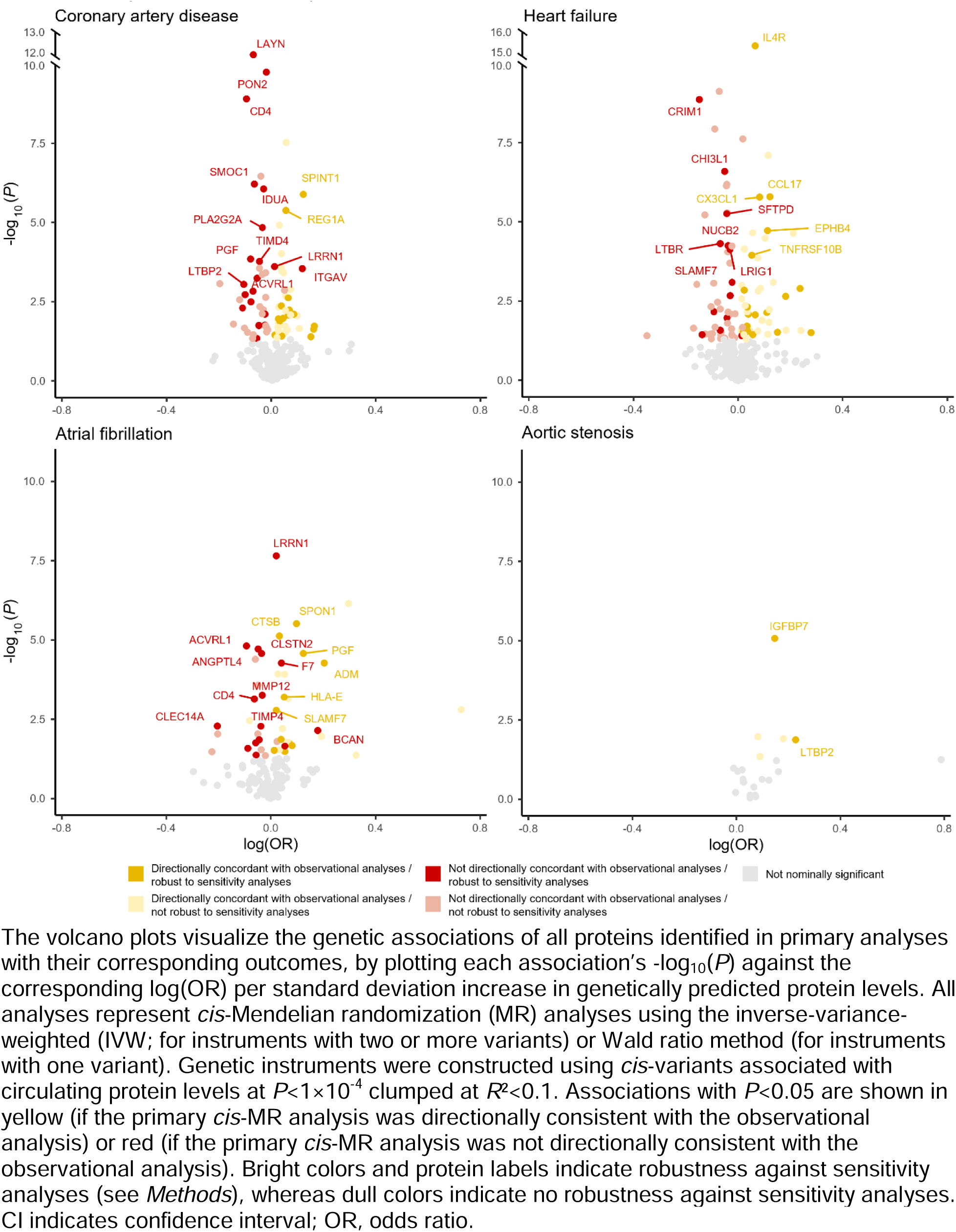
Associations of genetically predicted protein levels with coronary artery disease, heart failure, atrial fibrillation, and aortic stenosis.

Because proprotein convertase subtilisin/kexin type 9 (PCSK9) is an established causal biomarker and therapeutic target for coronary artery disease^22^, we used this protein as a positive control for our *cis*-MR analyses (despite PCSK9 not reaching Bonferroni significance for coronary artery disease in epidemiological analyses). Each SD increase in genetically predicted PCSK9 was associated with 1.28-fold odds of coronary artery disease (95%CI, 1.23-1.34; *P*=8.6×10^−30^), supporting a *cis*-MR strategy for the identification of potential causal protein-disease associations. The proteins with the strongest epidemiological associations did not generally show strong genetic associations with cardiac diseases. For instance, neither genetic associations for WFDC2 nor GDF15 reached nominal significance (**Supplementary Table 8**). Genetically predicted MMP12, which was among the most strongly associated proteins for incident coronary artery disease in epidemiological analyses but was only modestly associated with a protective effect on coronary artery disease risk in primary *cis*-MR analyses (OR, 0.98 [95%CI, 0.96-0.99] per SD; *P*=0.002).

Among all robust *cis*-MR analyses, the strongest genetic association was observed for the interleukin-4 receptor (IL4R), which was associated with 1.07-fold odds of heart failure (95%CI, 1.05-1.08; *P*=3.9×10^−16^) per SD of genetically predicted levels, consistent with previous work implicating heightened interleukin-4 signaling in profibrotic myocardial remodeling.^23^ The Kunitz-type protease inhibitor 1 (SPINT1; also known as hepatocyte growth factor activator inhibitor type 1) had the strongest directionally concordant and robust genetic association for coronary artery disease (OR, 1.13 [95%CI, 1.07-1.18] per SD; *P*=1.3×10^−6^). The senescence– and fibrosis-related insulin-like growth factor binding protein 7 (IGFBP7) was the most significant protein for aortic stenosis (OR, 1.10 [95%CI, 1.05-1.14] per SD; *P*=8.6×10^−6^). Spondin-1 (SPON1; OR, 1.10 [95%CI, 1.06-1.15]; *P*=3.0×10^−6^) and adrenomedullin (ADM; OR, 1.23 [95%CI, 1.11-1.35]; *P*=5.4×10^−5^) were among the strongest directionally concordant genetic associations for atrial fibrillation; notably, colocalization analyses suggested shared causal genetic variants between these two proteins and atrial fibrillation (posterior probability for shared causal variants [*H*_4_] >0.80). Colocalization analyses yielded inconclusive results for most other protein-disease associations (**Supplementary Table 13**).

### Sex-specific protein-disease associations

Although previous work suggested sex differences in the concentrations of cardiovascular biomarkers^24^, it remains unclear whether proteins assessed proteome-wide affect cardiovascular disease risk differently in men vs. women. Therefore, we tested the multivariable-adjusted associations of all 1,459 proteins with cardiac diseases in men (*n*=19,612) vs. women (*n*=24,701). A total of 467 protein-disease associations met the primary significance threshold (*P*<0.05/5,836) for men vs. 314 for women (**Supplementary Table 14**). Protein-disease associations (for all 1,459 tested biomarkers) showed strong correlation between sexes, indicated by a Pearson correlation coefficient (*r*) of 0.71 (**Supplementary** Fig. 4). The correlation between sexes was strongest for heart failure (*r*=0.79) whereas it was comparatively weaker for aortic stenosis (*r*=0.47).

We formally tested for sex interactions across all protein-disease associations reaching significance (*P*<0.05/5,836) in at least one sex (*n*=566) (**Fig. 4**, **Supplementary Table 14**). Six protein-disease associations had a Bonferroni-significant (*P*<0.05/566) sex-differential effect for atrial fibrillation, including T-cell surface glycoprotein CD1c (CD1C; HR_male_, 1.05 [95%CI, 0.99-1.11] vs. HR_female_, 0.90 [95%CI, 0.84-0.96] per SD; *P*_interaction_=6.9×10^−5^), cyclic ADP-ribose hydrolase (CD38; HR_male_, 1.01 [95%CI, 0.94-1.09] vs. HR_female_, 1.22 [95%CI, 1.12-1.34]; *P*_interaction_=3.3×10^−6^), cathepsin L2 (CTSV; HR_male_, 0.98 [95%CI, 0.22-1.05] vs. HR_female_, 0.87 [95%CI, 0.80-0.94]; *P*_interaction_=7.3×10^−5^), NT-proBNP (HR_male_, 1.69 [95%CI, 1.61-1.77] vs. HR_female_, 1.86 [95%CI, 1.74-1.99]; *P*_interaction_=3.7×10^−5^), paired immunoglobulin-like type 2 receptor β (PILRB; HR_male_,1.05 [95%CI, 0.99-1.11] vs. HR_female_, 1.20 [95%CI, 1.12-1.30]; *P*_interaction_=4.1×10^-5^), and WFDC2 (HR_male_, 1.20 [95%CI, 1.14-1.28] vs. HR_female_, 1.32 [95%CI, 1.23-1.42]; *P*_interaction_=7.7×10^−5^). We also observed a sex differential effect for chymotrypsin C (CTRC; HR_male_, 1.04 [95%CI, 0.99-1.09] vs. HR_female_, 0.88 [95%CI, 0.82-0.93]; *P*_interaction_=1.9×10^−5^) on coronary artery disease. For all of these associations, *P*-values were lower and effect sizes greater in women vs. men.

**Fig. 4.**
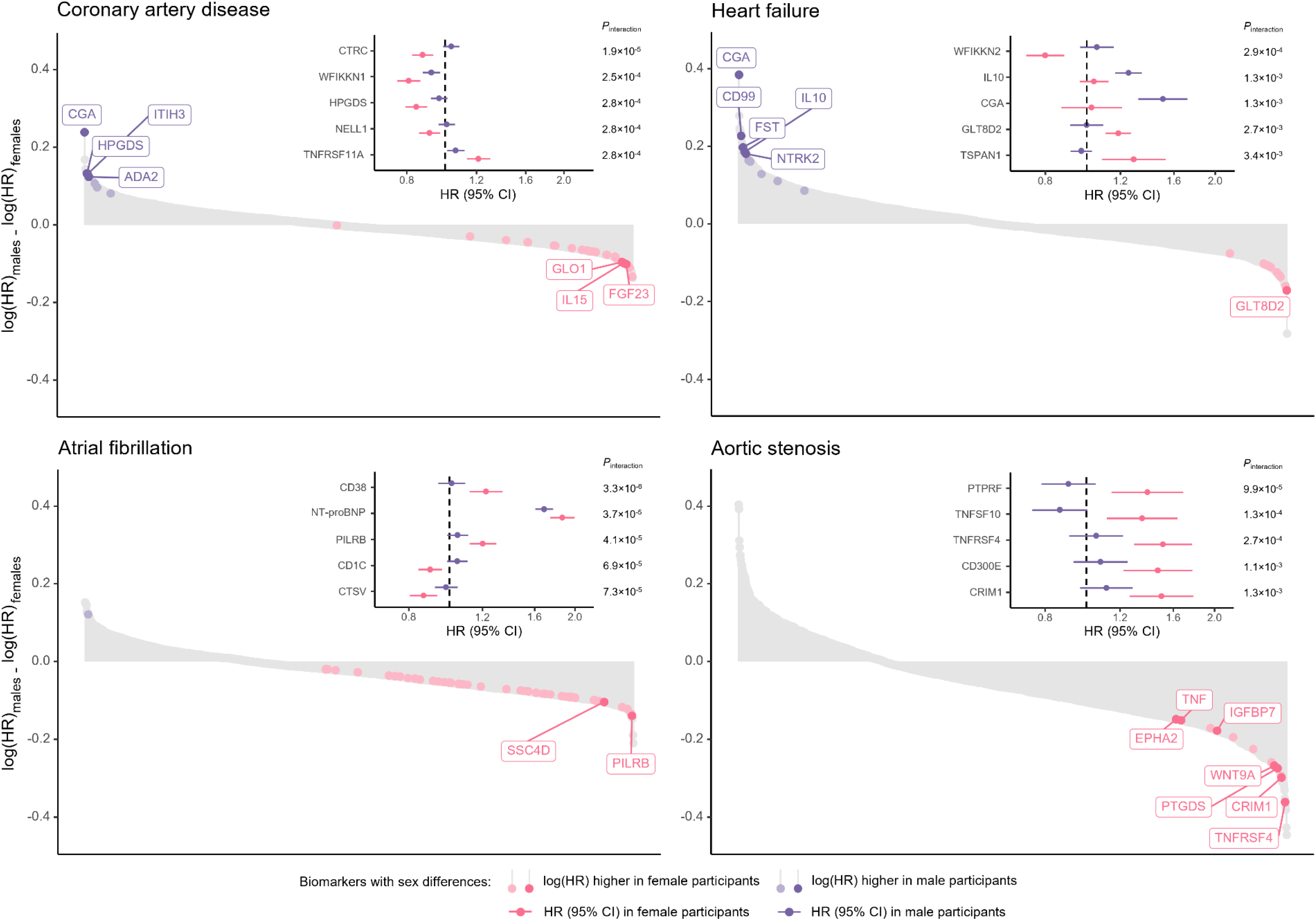
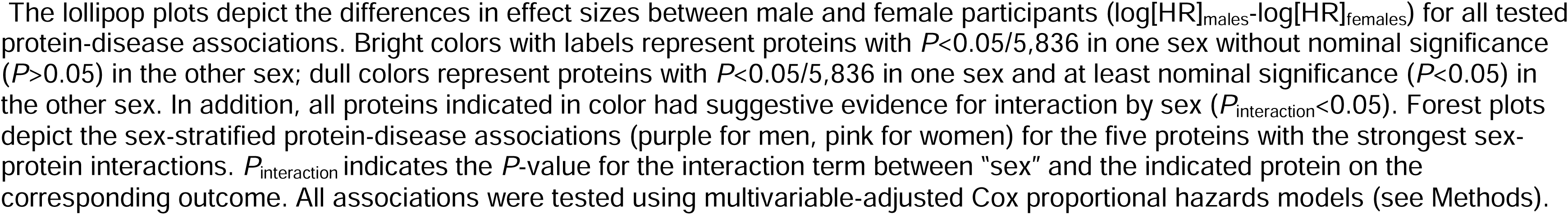
Sex-specific protein-disease associations and protein-by-sex interactions for coronary artery disease, heart failure, atrial fibrillation, and aortic stenosis.

### Protein-based prediction of cardiac diseases

We next derived and tested the predictive accuracy of protein-based risk scores in addition to clinical risk factors in the UKB-PPP. We constructed protein-based, clinical, and combined (using proteomic and clinical variables) risk scores in the training set (80%; *n*=35,450) using least absolute shrinkage and selection operator (LASSO) regression with 10-fold cross-validation. Protein-based risk scores included 64 proteins for coronary artery disease, 38 for heart failure, 92 for atrial fibrillation, and 21 for aortic stenosis (**Supplementary Table 15, Supplementary** Fig. 5). The prediction models’ highest-weighted biomarkers were largely overlapping with those showing the strongest associations in primary analyses.

Analyses in the testing cohort (20%; *n*=8,863) revealed that protein-based risk scores effectively stratified the risk of incident events across outcomes (**Fig. 5A-B**). The protein-based risk scores were strong independent predictors of incident events in multivariable-adjusted Cox regression models, with HRs of 1.83 (95%CI, 1.62-2.06; *P*=3.1×10^−23^) per SD increase for coronary artery disease, 2.33 (95%CI, 1.99-2.73; *P*=1.4×10^−25^) for heart failure, 2.22 (95%CI, 2.00-2.48; *P*=7.5×10^−48^) for atrial fibrillation, and 1.91 (95%CI, 1.39-2.64; *P*=7.5×10^−5^) for aortic stenosis. The top 10% vs. bottom 90% of protein-based risk scores was associated with HRs of 1.72 (95%CI, 1.38-2.15; *P*=1.8×10^−6^) for coronary artery disease, 3.62 (95%CI, 2.56-5.11; *P*=3.9×10^−13^) for heart failure, 4.02 (95%CI, 3.23-5.02; *P*=3.9×10^−35^) for atrial fibrillation, and 2.50 (95%CI, 1.31-4.77; *P*=5.5×10^−3^) for aortic stenosis.

**Fig. 5.**
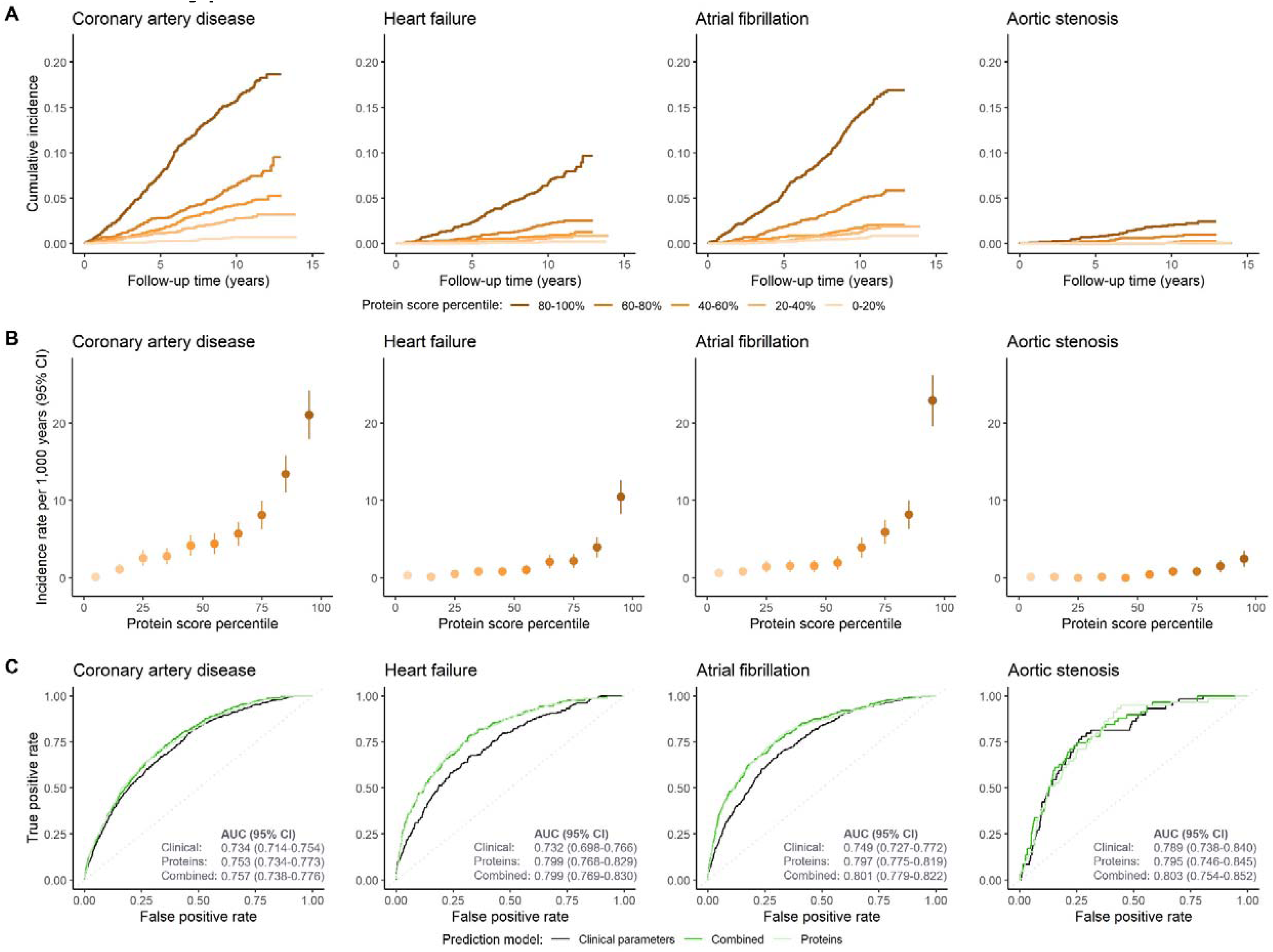
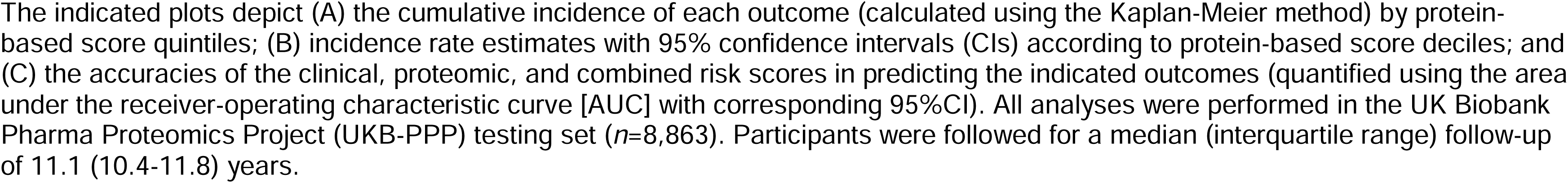
Risk (A-B) stratification and (C) prediction or incident coronary artery disease, heart failure, atrial fibrillation, and aortic stenosis by protein-based risk scores.

ROC curve analyses revealed that adding proteomic data improved the prediction of incident coronary artery disease, heart failure, and atrial fibrillation (**Fig. 5C**). The increment in predictive accuracy compared with the clinical model—quantified using the area under the ROC curve (AUC)—was most pronounced for atrial fibrillation (AUC, 0.801 [95%CI, 0.779-0.822] vs. 0.749 [95%CI, 0.727-0.772]; DeLong test: *P*=2.0×10^−10^) and heart failure (AUC, 0.799 [95%CI, 0.769-0.830] vs. 0.732 [95%CI, 0.698-0.766]; *P*=1.7×10^−6^), followed by coronary artery disease (AUC, 0.757 [95%CI, 0.738-0.776] vs. 0.734 [95%CI, 0.714-0.754]; *P*=1.4×10^−4^). There was no statistically significant difference for aortic stenosis (AUC, 0.803 [95%CI, 0.754-0.852] vs. 0.789 [95%CI, 0.738-0.840]; *P*=0.35).

Given the disproportionately high weights for NT-proBNP in the protein-based risk scores for atrial fibrillation and heart failure (**Supplementary Table 15**), we further evaluated the performance of models including NT-proBNP alone vs. those incorporating all other biomarkers in predicting these outcomes. We also excluded NPPB from the latter set of protein-based risk scores since NPPB and NT-proBNP are encoded by the same gene and released in the circulation in equimolar quantities^25^. Compared with the score based on clinical factors alone (0.749 [95%CI, 0.727-0.772]), inclusion of NT-proBNP significantly improved the prediction of atrial fibrillation (AUC, 0.788 [95%CI, 0.766-0.811]; *P*=1.1×10^−6^), resulting in a greater increment in predictive accuracy than the score incorporating all proteins other than NT-proBNP and NPPB (AUC, 0.777 [95%CI, 0.756-0.799]; *P*=9.2×10^−7^) (**Supplementary** Fig. 6). In contrast, for heart failure, the score incorporating all proteins except NT-proBNP and NPPB was associated with a greater improvement in predictive accuracy vs. the clinical score (AUC, 0.786 [95%CI, 0.754-0.818] vs. 0.732 [95%CI, 0.698-0.766]; *P*=1.4×10^−5^) than the score incorporating NT-proBNP alone (AUC, 0.756 [95%CI, 0.722-0.790]; *P*=0.07) (**Supplementary** Fig. 7).

## Discussion

In a population-based cohort of ∼45,000 middle-aged adults with circulating protein measurements and longitudinal follow-up, we characterized the proteomic architecture of incident coronary artery disease, heart failure, atrial fibrillation, and aortic stenosis. We identified 820 significant protein-disease associations with important roles (potentially mediating or marking disease presence) for natriuretic peptides (e.g., NT-proBNP), inflammatory mediators (e.g., MMP12), and apoptosis-related factors (e.g., GDF15) as predictors of cardiac diseases. Genetic analyses suggested causal or mediating roles—either protective or deleterious—for a substantial proportion of biomarkers identified in observational analyses. Sex-based analyses suggested generally preserved associations between men and women, albeit with varying weights of prediction including several biomarkers with strong sex interactions. Finally, we constructed sparse protein-based risk scores that improved the prediction of cardiac disease development in the general population. Our findings provide novel insights into the biology of cardiac diseases, with implications for the prediction of incident cardiovascular diagnoses, and with the potential for targeted prevention and treatment of these conditions.

The findings from this study offer insights into potential causal roles of proteins associated with incident cardiac diseases. We found that ∼6% of proteins identified in primary analyses (and tested in *cis*-MR analyses) had putative causal associations that were directionally discordant with those derived from epidemiological models (i.e., primary analyses). However, we also identified many proteins (another ∼6%) with genetic associations that were robust across sensitivity analyses yet discordant with epidemiological estimates. By systematically integrating observational and genetic data, our study corroborates and extends previous studies reporting similar discrepancies between genetic and epidemiological associations for selected proteins^15,26^. For instance, consistent with prior research^15,26^, primary *cis*-MR analyses suggested a protective effect of genetically predicted MMP12 on coronary artery disease risk, despite observational analyses indicating strong associations of higher MMP12 levels with the same outcome. Similarly, higher measured but lower genetically predicted levels of layilin (LAYN)—a relatively poorly understood protein that binds hyaluronic acid and has roles in cellular adhesion and immunity^27^—were strongly and robustly associated with increased coronary artery disease risk. Whether these seemingly discordant observations reflect inherent differences between disease onset versus progression or compensatory response to subclinical disease requires further investigation. Nevertheless, several proteins had consistent genetic and observational effects. One such example was SPON1, for which higher levels (both measured and genetically predicted) were associated with increased atrial fibrillation risk. SPON1 is an extracellular protein expressed in tissues such as the heart and brain that has been implicated in Alzheimer’s dementia through its role in amyloid-β precursor protein processing^28^. Previous protein-focused analyses in patients with heart failure showed that the presence of atrial fibrillation was associated with activation of amyloid-β-related pathways, with SPON1 as one of the most strongly upregulated proteins in those with atrial fibrillation^29^. These data, together with colocalization findings indicating shared causal variants for SPON1 and atrial fibrillation, collectively suggest that SPON1 not only marks presence of a pro-arrhythmic substrate, but could also represent an upstream therapeutic target for preventing and/or treating atrial fibrillation. Given the paucity of identified robust biomarkers mediating atrial fibrillation risk, more data are needed regarding SPON1 and its role in arrhythmogenesis.

Biomarkers identified in proteomic analyses are often markers of already established disease, rather than mediators of disease biology. In this regard, our analyses identified several inflammation– and apoptosis-related proteins as strong predictors marking risk for cardiac disease but were unlikely causal biomarkers. WFDC2 (also known as HE4)—a profibrotic protease inhibitor with a potential role in natural immunity^17^—emerged as the strongest proteomic predictor of heart failure. Previous research in hospitalized heart failure patients demonstrated associations of circulating WFDC2 with disease severity as well as kidney function^30^. As WFDC2 is expressed exclusively in non-cardiovascular tissues such as the respiratory tract, male and female genitourinary system, and kidneys^31^, it is likely that the strong associations of WFDC2 with cardiac outcomes stem from peripheral organ responses rather than indicating direct cardiac dysfunction or vascular damage. Similarly, GDF15—another pleiotropic protein expressed across multiple organ systems^32^—was the strongest biomarker for coronary artery disease. As a member of the transforming growth factor-β (TGF-β) superfamily, GDF15 is upregulated in response to external stressors (e.g., inflammation, hypoxia, and oxidative stress) and is believed to reflect the cumulative impact of both acute and chronic exposure to cellular stressors^32^. Recent data suggest GDF15 as an independent prognostic biomarker for individuals with established atherosclerotic cardiovascular disease^33^. The present study builds upon this work, extending the applicability of WFDC2 and GDF15 as cardiac biomarkers to the general population. Nevertheless, *cis*-MR analyses found no evidence of causality in the associations of these proteins with cardiac diseases. These findings collectively suggest that inflammation– and apoptosis-related biomarkers such as WFDC2 and GDF15 represent early disease markers without causal involvement in the pathogenesis of cardiac diseases, consistent with their pleiotropic and non-specific effects in response to tissue damage across organs. Much the same may be said about the natriuretic peptides.

In this regard, findings of this study corroborate the role of natriuretic peptides as important cardiac biomarkers, grossly up-regulated in those with already prevalent but sub-clinical cardiovascular disease states. In addition to the established association of natriuretic peptide elevation with so-called “pre-heart failure”^34^, the result of this study support the use of NT-proBNP to guide atrial fibrillation screening. NT-proBNP is the N-terminal fragment of proBNP (i.e., the precursor of biologically active NPPB, also known as BNP) and is primarily released by ventricular and/or atrial cardiomyocytes in response to elevated myocardial wall tension^25^. Proteomic association analyses identified NT-proBNP as one of the strongest predictors of heart failure, consistent with its routine clinical use in diagnosing and risk-stratifying this condition^25^. Despite its widespread use in the setting of heart failure (where it was a strong marker), the results of this study suggest that NT-proBNP’s strongest disease associations were observed for atrial fibrillation; ROC curve analyses also revealed that NT-proBNP contributed more to the prediction of atrial fibrillation than to that of heart failure. These results extend prior work demonstrating strong associations of circulating NT-proBNP with incident atrial fibrillation^35^ and align with recent data from the LOOP trial suggesting that individuals with elevated NT-proBNP levels may derive more clinical benefit from atrial fibrillation screening than those with lower levels^36^. Collectively, these findings provide support for the use of NT-proBNP to guide atrial fibrillation screening in the general population.

Another important finding from this study was evidence for biological sex differences underlying cardiac disease risk in men and women. The strongest sex interaction across all tested proteins was observed for CD38, which was significantly more strongly associated with incident atrial fibrillation in female than in male participants. CD38 is a glycoprotein expressed across various immune cells including lymphocytes and plasma cells^37^. Previous research suggests that CD38 is causally implicated in autoimmune diseases such as rheumatoid arthritis and systemic lupus erythematosus^37,38^. As a history of autoimmune diseases represents a risk factor for atrial fibrillation that affects women more strongly than men^39^, it could be possible that the observed sex differences for CD38 reflect a more important role for immunity-related pathways in women vs. men. Furthermore, some of the largest differences in protein-disease associations between sexes were observed for aortic stenosis. Prior histological work in aortic stenosis patients suggests distinct tissue composition differences between men and women, with women showing less valvular calcification but more fibrosis than men^40^. Building upon these findings, we identified several sex-specific senescence-associated biomarkers (e.g., IGFBP7 and TNF) associated with incident aortic stenosis in female, but not male, participants. Notably, *cis*-MR analyses indicated a potential causal effect of IGFBP7 on aortic stenosis risk. IGFBP7 plays a role in the senescence-associated secretory phenotype, promoting cell cycle arrest upon tissue damage and leading to apoptosis and tissue fibrosis^41^. These findings support an important role for fibrotic valve remodeling in aortic stenosis, particularly among women, identifying IGFBP7 as a mediator and potential therapeutic target in this population.

While this study benefits from a large sample size and the use of state-of-the-art proteomic profiling methods, findings must be interpreted in the context of limitations. First, the strength and quantity of protein-disease associations for each outcome were influenced by statistical power and, consequently, the number of cases per outcome. Conditions with lower incidence rates during follow-up (such as aortic stenosis) had fewer proteomic associations. Second, the study population was predominantly white, precluding generalization to other races/ethnicities. Third, any causal inference using MR relies on the validity of the underlying instrumental variable assumptions. This study utilized a robust *cis*-MR framework (facilitating the adherence to these assumptions^21,42^) and tested the robustness of the genetic associations through many sensitivity analyses, suggesting potential causal roles for the identified proteins. Nevertheless, prioritized therapeutic targets remain to be evaluated in animal experiments and eventually human trials. Fourth, not all proteins identified in primary analyses had strong *cis*-pQTLs, precluding adequate *cis*-MR analyses for these proteins. Additionally, genetic instrument strength varied across proteins. Instruments with more variants have greater power to detect statistically significant genetic protein-disease associations, potentially leading to an underestimation of associations for instruments with fewer variants. Nevertheless, we minimized type II error by adopting a lenient *P*-value threshold (*P*<0.05) to indicate statistical significance for primary *cis*-MR analyses and prioritizing genetic protein-disease associations that were robust to many sensitivity analyses.

Leveraging a population-based cohort of ∼45,000 participants with comprehensive genotypic, proteomic, and longitudinal clinical profiling, this study characterized the circulating proteome associated with incident coronary artery disease, heart failure, atrial fibrillation, and aortic stenosis. The study findings support new applications for established biomarkers (e.g., atrial fibrillation surveillance using NT-proBNP) and identify strong and potentially useful predictors of cardiac diseases (e.g., WFDC2 for heart failure). Additionally, this study prioritized potential therapeutic targets within the circulating proteome and elucidated biological sex differences. These results demonstrate the potential of proteomics in refining cardiovascular risk assessment and lay a foundation for future investigations to uncover novel disease mechanisms and assess the clinical utility of protein-based prevention strategies for cardiac diseases.

## Methods

### Study design and participants

The study design is illustrated in **Fig. 1**. The UK Biobank is a population-based cohort of ∼500,000 volunteers aged 40-69 years at the time of study enrolment, recruited from 22 assessment centers across the United Kingdom during 2006-2010.^43^ At enrolment, participants provided informed consent; underwent physical examination; provided details on sociodemographic characteristics, lifestyle factors, medical history, and medication use; and donated blood samples. Follow-up for incident events occurred via linkage to electronic health records through March 2020.

The UK Biobank Pharma Proteomics Project (UKB-PPP) is a precompetitive consortium of 13 biopharmaceutical companies funding the generation of blood-based proteomic data in a subset of UK Biobank participants.^16,43^ Upon release, the sponsors have no direct role in research activities of these features as is the case for the present work. The UKB-PPP includes 54,306 participants, of whom 46,673 (85.9%) were randomly selected from baseline, 6,385 (11.8%) were preselected by UKB-PPP consortium members based on certain characteristics of interest (e.g., disease status or genetic ancestry), and 1,268 (2.3%) were selected because they attended multiple visits of the COVID-19 case-control imaging study.^16^ We considered 52,705 participants with baseline proteomic data passing quality control for inclusion in the present study (**Fig. 1**). Participants were excluded if they had missing data for >10% of assay measurements or if they had missing data on self-reported race/ethnicity or genetic ancestry. We also excluded individuals inferred to be related (closer than third degree; kinship coefficient >0.0884) and those with self-reported or physician-ascertained coronary artery disease, heart failure, atrial fibrillation, or aortic stenosis at baseline (see below for disease definitions).

The UK Biobank was approved by the North West Multi-center Research Ethics Committee. All analyses were conducted under UK Biobank application number 7089. The Mass General Brigham Institutional Review Board approved the secondary use of these data.

### Protein measurements and proteomic data processing

A detailed overview of the protein measurement strategy and proteomic data processing steps in the UKB-PPP can be found in the **Supplementary Note** and has been described previously.^45^ In brief, blood samples donated by UKB-PPP study participants at study baseline underwent proteomic profiling using the *Olink Explore 1536* platform (Olink Proteomics, Inc; Waltham, MA), which uses proximity extension assay technology to measure 1,472 protein analytes across four different panels (the *Cardiometabolic*, *Inflammation*, *Neurology*, and *Oncology* panels) representing 1,463 unique proteins (**Supplementary Table 1**). We excluded proteins with >10% missingness in the final study cohort (cathepsin S, nucleophosmin, tumor-associated calcium signal transducer 2, and procollagen C-endopeptidase enhancer 1; **Supplementary Table 2**) and imputed the remaining 1.1% of missing protein values using *K*-nearest neighbors (*K*=10) as done previously.^44^ All 1,459 protein markers underwent *Z*-score transformation prior to analysis.

### Outcome ascertainment

Follow-up for incident outcomes occurred through linkage to national health records until March 2020. Incident events were defined by the occurrence of (1) at least one qualifying ICD-9 or ICD-10 code for a corresponding in-or outpatient diagnosis (as either a primary or secondary disease diagnosis); or (2) at least one *Office of Population Censuses and Surveys Classification of Surgical Operations and Procedures* (OPCS) code for a qualifying procedure (e.g., coronary artery revascularization for coronary artery disease). The specific codes used to define each outcome are listed in **Supplementary Table 16** and were used as done previously.^46,47^

### Proteomic association analyses

Primary analyses tested the associations of circulating protein levels with incident cardiac events using Cox proportional hazards models adjusted for age, age², sex, self-reported race/ethnicity, the first ten principal components of genetic ancestry, smoking, normalized Townsend deprivation index, BMI, systolic blood pressure, antihypertensive medication use, total cholesterol, HDL cholesterol, cholesterol-lowering medication use, serum creatinine (as a measure of kidney function), and prevalent type 2 diabetes. In addition, to increase the specificity of the detected protein associations for a given disease (e.g., coronary artery disease), we included the other cardiac outcomes (e.g., heart failure, atrial fibrillation, and aortic stenosis) as time-varying covariates using the *tmerge()* function in R (*survival* package).^48^ A detailed description of the study covariates can be found in the **Supplementary Note**. Bonferroni-corrected *P*<0.05/5,836 (0.05/[1,459×4] or ∼8.6×10^−6^) indicated statistical significance for the primary analyses.

### Pathway enrichment analyses

Pathway enrichment analyses evaluated whether certain protein groups representing biologically distinct pathways were disproportionately up– or downregulated in individuals with incident cardiac events. Top biological functions, molecular pathways, and cellular components were queried for each outcome using the *Gene Ontology* resource^18^ via *Enrichr.*^19^ Enrichment tests were performed against a background gene set including the genes corresponding to all 1,459 proteins tested in primary analyses. Gene sets with a false discovery rate-adjusted *P*<0.05 were considered statistically significant.

### Cis-Mendelian randomization analyses

We performed two-sample MR analyses to explore the causal roles of proteins that were statistically significantly associated with one or more cardiac outcomes in epidemiological models (i.e., primary analyses). These analyses tested the associations of protein quantitative trait loci (pQTLs; genetic variants associated with circulating protein levels) with coronary artery disease, heart failure, atrial fibrillation, and aortic stenosis. We obtained pQTL data from 35,571 UKB-PPP participants who had their circulating proteomes profiled using the *Olink Explore 1536* platform.^16^ FinnGen (freeze 9; https://r9.finngen.fi/) was used for genetic association data for coronary artery disease (cases/total participants: *n*=43,518/377,277), heart failure (*n*=27,304/377,277), atrial fibrillation/flutter (*n*=45,766/237,690), and operated calcific aortic stenosis (*n*=9,153/377,277). All genetic data were derived from individuals of European ancestry, and there was no overlap between the exposure and outcome study cohorts.

Because the use of *cis*-pQTLs (pQTLs that map near the protein-encoding gene) facilitates adherence to MR assumptions,^21,26^ we only used region-wide significant, largely independent *cis*-variants to construct genetic instruments (*P*<5×10^−6^, *R*²<0.1, ±200 kilobases). All proteins with at least one valid *cis*-pQTL were tested. We used the inverse-variance-weighted (IVW) method for genetic instruments with more than one *cis*-pQTL and the Wald ratio estimator for instruments with only one *cis*-pQTL. IVW estimates were adjusted for residual correlation between genetic variants.^49,50^ Sensitivity analyses were performed for all genetic protein-disease associations with at least nominal significance (unadjusted *P*<0.05) in primary *cis*-MR analyses. These sensitivity analyses tested whether the observed genetic associations were robust to (1) different instrument construction parameters (*R*²<0.001/<0.01/<0.1/<0.2 and *P*<5×10^−4^/*<*5×10^-6^/<5×10^−8^); (2) MR-Egger (which tests for horizontal pleiotropy); (3) one-sample MR (in an independent UKB sample); and (4) multivariable-adjusted MR adjusting for genetic instruments of proteins which may also exert *trans*-pQTL effects.

Genetic associations were considered robust if the effect estimates were directionally consistent across all primary and sensitivity analyses and MR-Egger suggested no horizontal pleiotropy. Additional details of the MR methodology can be found in the **Supplementary Note**.

### Protein-based prediction models

We constructed protein-based risk scores to predict incident cardiac events in the UKB-PPP. We created three risk scores for each cardiac outcome using logistic least absolute shrinkage and selection operator (LASSO) regression, including (1) a score based on clinical risk factors, (2) a score based on circulating proteins, and (3) a combined score (i.e., using clinical risk factors and circulating proteins). The clinically evaluable variables used as covariates in primary analyses (age, sex, self-reported race/ethnicity, smoking, BMI, systolic blood pressure, antihypertensive medication use, total cholesterol, HDL cholesterol, cholesterol-lowering medication use, serum creatinine, and type 2 diabetes) were fed into LASSO models for the clinical risk scores. Circulating levels of the 1,459 proteins tested in primary analyses were fed into LASSO models for the protein-based risk scores.

The study cohort was randomly divided into a training (80%; *n*=35,450) and testing (20%; *n*=8,863) set. All risk scores were constructed in the training set using LASSO regression with 10-fold cross-validation. The performance of each prediction model was evaluated by receiver-operating characteristic (ROC) curve analysis. Additional information on the construction and evaluation of all prediction models can be found in the **Supplementary Note**.

## Acknowledgements

All UK Biobank analyses were performed under application N° 7089.

Acknowledgements

A.S. is supported by the Belgian American Educational Foundation. J.L.J. is supported in part by the Adolph M. Hutter MD Professorship. M.C.H. is supported by the U.S. NHLBI (K08HL166687) and AHA (940166, 979465). P.N. is supported by the Paul & Phyllis Fireman Endowed Chair in Vascular Medicine from the Massachusetts General Hospital and the U.S. NHLBI (R01HL127564).

## Ethics declarations

J.L.J. is a Trustee of the American College of Cardiology and reports board membership of Imbria Pharmaceuticals; grant support from Abbott Diagnostics, Applied Therapeutics, AstraZeneca, Bayer, HeartFlow, Innolife and Roche; previous consulting income from Abbott Diagnostics, AstraZeneca, Bayer, Beckman-Coulter, Jana Care, Janssen, Novartis, Quidel, Roche Diagnostics and Siemens; clinical endpoint committee/data safety monitoring board membership for Abbott, AbbVie, CVRx, Pfizer and Takeda. M.C.H. reports consulting fees from CRISPR Therapeutics and Comanche Biopharma; advisory board service for Miga Health; and grant support from Genentech. P.N. reports research grants from Allelica, Amgen, Apple, Boston Scientific, Genentech / Roche, and Novartis; personal fees from Allelica, Apple, AstraZeneca, Blackstone Life Sciences, Creative Education Concepts, CRISPR Therapeutics, Eli Lilly & Co, Foresite Labs, Genentech / Roche, GV, HeartFlow, Magnet Biomedicine, Merck, and Novartis; scientific advisory board membership of Esperion Therapeutics, Preciseli, and TenSixteen Bio; scientific co-foundership of TenSixteen Bio, equity in MyOme, Preciseli, and TenSixteen Bio; and spousal employment at Vertex Pharmaceuticals (all unrelated to the present work).

## Data availability

Data supporting the results of the present study are available from the UK Biobank (https://www.ukbiobank.ac.uk/enable-your-research/apply-for-access) to bona fide researchers with institutional review board and UK Biobank approval. These analyses were performed using the UK Biobank resource under application number 7089. The secondary use of these data was approved by the Mass General Brigham institutional review board. Pathway enrichment analyses were performed using the Gene Ontology resource via Enrichr (https://maayanlab.cloud/Enrichr/). The UK Biobank Pharma Proteomics Project was used for genetic association data for circulating proteins (i.e., protein quantitative trait locus data) through Synapse (https://doi.org/10.7303/syn51364943). FinnGen was used for genetic association data for coronary artery disease, heart failure, atrial fibrillation/flutter, and operated calcific aortic stenosis (freeze 9; https://r9.finngen.fi/).

## Code availability

Code used for the main analyses of this study can be accessed at https://github.com/aschuerm/ukbppp_cardiac_diseases.

## Inclusion and ethics

Inclusion and ethics standards have been reviewed where applicable.

