## Supplementary material for "Integrative proteomic analyses across common cardiac diseases yield new mechanistic insights and enhanced prediction": Suppl. Note

### Supplementary Methods

#### Protein measurements, quality control, and data normalization

Blood samples donated by UKB-PPP study participants underwent proteomic profiling using the *Olink Explore 1536* platform (Olink Proteomics, Inc; Waltham, MA), which measures 1,472 protein analytes across four different panels (the *Cardiometabolic*, *Inflammation*, *Neurology*, and *Oncology* panels) representing 1,463 unique proteins (**Supplementary Table 1**). Details on this platform have been described previously.^1^ In brief, Olink uses proximity extension assay technology, whereby antibody pairs with conjugated oligonucleotides bind their target proteins in a pairwise manner. When an antibody pair has bound its target, complementary oligonucleotides undergo hybridization and, subsequently, extension by DNA polymerase. These DNA sequences—or tags—are then amplified through polymerase chain reaction amplification, which can be quantified using next-generation sequencing. For each assay and each sample, normalized protein expression values are calculated as the log_2_-transformed ratio of sequence read counts to the counts of the extension control, corrected for plate and batch effects. Additional details on data normalization and quality control have been described previously.^2,3^

For proteins that were measured by multiple panels (TNF, IL6, and CXCL8), we only evaluated data from the panel with the highest detectability per protein and, if necessary, the largest number of protein measurements exceeding the respective limit of detection (the *Cardiometabolic* panel for TNF and *Oncology* panel for IL6 and CXCL8). We further excluded proteins with >10% missingness in the final study cohort (CTSS and NPM1 from the *Neurology* panel, PCOLCE from the *Cardiometabolic* panel, and TACSTD2 from the *Oncology* panel; **Supplementary Table 2**) and imputed the remaining 1.1% of missing protein values using *K*-nearest neighbors (*K*=10) via the *impute.knn()* function (*impute* package^4^ in R) as done previously.^2^ The remaining 1,459 protein markers underwent *Z*-score transformation prior to analysis.

#### Covariate ascertainment

Demographic characteristics, medical history, medication use, and health behaviors were systematically assessed upon enrolment in the UK Biobank. Self-reported-race/ethnicity was collected at baseline and used as a binary variable (White vs. non-White) in analysis. Smoking was dichotomized as ever (i.e., current or past) smoking vs. no history of smoking. Type 2 diabetes was defined by self-report or qualifying *International Classification of Diseases* (ICD) codes (**Supplementary Table 3**). The Townsend deprivation index—an area-level score that incorporates data on home ownership, automobile ownership, employment, and household overcrowding—was used as a composite measure of material deprivation.^5^ Townsend deprivation index scores were inverse-rank normalized and *Z*-score transformed before analysis.

Anthropometric data, physical measurements, and blood samples were obtained by trained study staff.^6^ Body mass index (BMI) was calculated from standing height and weight measured at baseline. After a five-minute period of seated rest, blood pressure was measured using an electronic monitor (Omron 705 IT, OMRON Healthcare) on two separate occasions with a one-minute interval in between; the mean was calculated and used for analysis when both measurements were available. Total cholesterol, high-density lipoprotein (HDL) cholesterol, and creatinine concentrations were quantified in baseline blood samples (AU5800, Beckman Coulter).

Missing values for BMI (missing for *n*=717; 1.6%), systolic blood pressure (*n*=2,210; 5.0%), total cholesterol (*n*=1,995; 4.5%), HDL cholesterol (*n*=5,591; 12.6%), serum creatinine (*n*=2,010; 4.5%), and normalized Townsend deprivation index (*n*=53; 0.1%) were imputed using linear regression models incorporating sex, age, race/ethnicity, and the first ten principal components of genetic ancestry as predictors.

#### *Cis*-Mendelian randomization analyses

##### Main analyses

Because the use of *cis*-pQTLs (i.e., pQTLs that map near the protein-encoding gene) facilitates adherence to the assumptions of MR,^7,8^ we only used variants within a 200-kilobase range of the protein-encoding gene to construct our genetic instruments. We used a relaxed *P*-value threshold for instrument selection (*P*<5×10^-6^) relative to the conventional genome-wide threshold (*P*<5×10^-8^) to increase the number of genetic instruments since the *cis*-regions for the assayed proteins represent only a small fraction of the genome, are expected to be enriched for associations, and to optimize power. All *cis*-pQTLs with *P*<5×10^-6^ were clumped into largely independent loci (linkage disequilibrium *R*²<0.1) using PLINK.^9^ Linkage disequilibrium information was obtained from the European panel of phase 3 of the *1000 Genomes Project.*^10^

To minimize the risk of weak instrument bias, we only performed *cis*-MR analyses for genetic instruments with *F*-statistics >10. *F*-statistics were obtained by performing linear regression analyses of a protein’s genetic risk score (as the independent variable) against the measured levels of that protein (as the dependent variable) in the UKB-PPP. Genetic risk scores were calculated as weighted allele scores using the “clumping and thresholding” method, applying the same *P*-value and linkage disequilibrium *R*² thresholds as those used in our primary *cis*-MR analyses (*P*<5×10^-6^ and *R*²<0.1). All scores were calculated using genotype array data; for proteins where genetic risk score calculation failed, *F*-statistics were estimated using summary statistics as

$$\text{F }\text{= }\frac{\text{(}\text{n}\text{ }\text{- }\text{k}\text{ }\text{- 1)}}{\text{k}}\text{ × }\frac{(\text{R}^{\text{2}})}{\text{(1 - }\text{R}^{\text{2}})}$$

where *n* indicates the sample size of the original genome-wide association study, *k* the number of variants included in the genetic instrument, and *R*² the variance in the exposure explained by the genetic variants, as done previously.^11^

Depending on the number of *cis*-pQTLs included in a protein's genetic instrument, we used different MR methods to infer causal effects.^12^ The inverse-variance-weighted (IVW) method was used with fixed effects for genetic instruments with two to three *cis*-pQTLs and with multiplicative random effects for those with more than three *cis*-pQTLs. The Wald ratio estimator was used for genetic instruments with only one *cis*-pQTL. We adjusted for between-variant correlation structure in all IVW models to avoid inflated estimates caused by residual correlation, as described previously.^13,14^

##### Sensitivity analyses

Because it is routinely recommended to evaluate the robustness of *cis*-MR estimates using multiple sensitivity analyses,^15^ we performed additional analyses using different MR approaches and instrument selection parameters. First, we evaluated the possibility of reverse causation affecting our analyses by performing Steiger filtering to exclude variants explaining more variance in the outcome (i.e., cardiac diseases) than the exposure (i.e., circulating protein levels). Second, as *cis*-MR analyses often rely on pQTLs that may be residually correlated with each other, we carried out sensitivity analyses testing genetic instruments that were constructed using a range of linkage disequilibrium *R*² thresholds (*R*²<0.001, *R*²<0.01, *R*²<0.1, and *R*²<0.2). Third, because the primary genetic instruments were constructed using sub-genome-wide significant *cis*-pQTLs, we verified the robustness of our genetic associations against different *P*-value thresholds (*P*<5×10^-4^, *P*<5×10^-6^, and *P*<5×10^-8^). Fourth, we calculated effect estimates using the MR-Egger method to account for horizontal pleiotropy (i.e., effects of the genetic instruments on the outcome through pathways other than the exposure of interest). Fifth, we performed one-sample *cis*-MR analyses to test the associations of the prioritized proteins’ genetic risk scores with cardiac diseases in an external UKB sample (cfr. below). Sixth, to account for the possibility that a certain protein’s genetic instrument could affect the outcomes through one or more other proteins, we calculated multivariable-adjusted *cis*-MR estimates that were adjusted for the genetic instruments of all prioritized proteins significantly associated with the tested protein’s genetic risk score (cfr. below).

Genetic risk scores were calculated from genetic association data from the UKB-PPP as weighted allele scores using the “clumping and thresholding” method, applying the same *P*-value and linkage disequilibrium *R*² thresholds as those used in the primary *cis*-MR analyses (*P*<5×10^-6^ and *R*²<0.1). One-sample *cis*-MR was performed in UKB participants who were not included in the UKB-PPP, were free of cardiac diseases at baseline, and had no missing covariates (*N*=407,230). Associations of the circulating proteins’ genetic risk scores with cardiac outcomes were tested using Cox regression models adjusted for age, age², sex, race/ethnicity, and the first ten principal components of genetic ancestry. In addition, we interrogated the possibility that a certain protein’s genetic instrument was also associated with other proteins’ circulating levels (i.e., proteins with shared pQTLs). To investigate this, we tested the associations of genetic risk scores for all proteins surviving upstream sensitivity analyses with all proteins with putative causal associations in the primary *cis*-MR analyses. Linear regression models adjusted for age, age², sex, race/ethnicity, and the first ten principal components of genetic ancestry were employed for these analyses. For proteins with genetic instruments that were significantly associated with one or more other proteins (i.e., “correlated proteins”), we then calculated multivariable-adjusted *cis*-MR estimates in the independent UKB sample (*N*=407,230) using Cox regression models adjusted for age, age², sex, race/ethnicity, the first ten principal components of genetic ancestry, and the genetic risk scores of all “correlated” proteins.

Sensitivity analyses were performed for all genetic protein-disease associations with at least nominal significance (unadjusted *P*<0.05) in primary *cis*-MR analysis. Genetic associations were considered robust if (1) the effect estimates were directionally consistent across all primary and sensitivity analyses and (2) MR-Egger suggested no horizontal pleiotropy (i.e., *P*≥0.05 for the intercept test or *P*<0.05 for the intercept test with *P*<0.05 for the causal test). MR analyses were performed using the *TwoSampleMR* and *MendelianRandomization* packages in R.^16,17^

#### Colocalization analyses

We performed colocalization analyses to test for shared causal variants between the prioritized proteins' *cis* loci (from the UKB-PPP) and corresponding cardiac outcomes (from FinnGen). Analyses considered all variants that were present in the protein and outcome summary statistics within ±200 kilobases of each biomarker’s protein-encoding region. Colocalization analyses were performed using the *coloc.abf()* function (*coloc* package^18^ in R). All colocalization analysis results were expressed as test statistics representing the posterior probabilities of five hypotheses: *H*_0_, neither trait has an association with a genetic variant in the region; *H*_1_, only the indicated protein has an association with a genetic variant in the region; *H*_2_, only the indicated cardiac disease has an association with a genetic variant in the region; *H*_3_, both traits are associated but with different causal variants; and *H*_4_, both traits are associated and share a single causal variant. A posterior probability for *H*_4_ >0.80 indicated strong colocalization evidence.

#### Sex-stratified association analyses

Sex-stratified analyses tested the associations of circulating protein levels with incident coronary artery disease, heart failure, atrial fibrillation, and aortic stenosis in self-reported female and male participants separately. These analyses were performed using Cox proportional hazards models adjusted for age, age², self-reported race/ethnicity, the first ten principal components of genetic ancestry, smoking, normalized Townsend deprivation index, BMI, systolic blood pressure, antihypertensive medication use, total cholesterol, HDL cholesterol, cholesterol-lowering medication use, serum creatinine, and prevalent type 2 diabetes. Coronary artery disease, heart failure, atrial fibrillation, and aortic stenosis were included as time-varying covariates. The difference in effect size for the protein-disease associations was quantified by substracting the natural logarithm of the hazard ratio (HR) in females from the natural logarithm of the HR in males (log[HR]_males_-log[HR]_females_).

We tested all protein-disease association reaching significance (*P*<0.05/5,836) in at least one sex for protein-by-sex interactions. These analyses were performed by fitting an interaction term (sex × circulating protein levels) in Cox proportional hazards models adjusted for sex, age, age², self-reported race/ethnicity, the first ten principal components of genetic ancestry, smoking, normalized Townsend deprivation index, BMI, systolic blood pressure, antihypertensive medication use, total cholesterol, HDL cholesterol, cholesterol-lowering medication use, serum creatinine, prevalent type 2 diabetes, and circulating levels of the tested protein.

#### Construction of protein-based prediction models

All clinical, proteomic, and combined prediction scores were constructed using the least absolute shrinkage and selection operator (LASSO) model for variable selection and regularization. In brief, LASSO is a regularized regression method that selects informative variables (e.g., proteins or clinical risk factors) from high-dimensional and correlated datasets while shrinking the regression coefficients of less informative variables to zero. We used 10-fold cross-validation to tune the regularization parameter (*λ*; the parameter that controls the strength of shrinkage and variable selection) for each LASSO model. During the cross-validation procedure, multiple LASSO models are iteratively constructed for each set of predictors (i.e., clinical risk factors, proteins, or both) using different values for *λ*, with each *λ* corresponding to a certain number of variables included in the prediction model. A higher *λ* value corresponds to fewer predictive variables in the regression model. The accuracy of each LASSO model (with its respective *λ* value) was quantified using the area under the receiver-operating characteristic curve (AUC).

We used the “one standard error rule” to determine the optimal *λ* for all proteomic and combined prediction models. This approach reduces the complexity of prediction models that are derived from high-dimensional datasets by selecting the largest *λ* (which corresponds to the smallest number of predictive covariates) for which the AUC is within one standard error of the highest AUC value during the cross-validation process. For models based solely on clinical risk factors, the *λ* corresponding to the highest AUC was used, considering that these risk scores were derived from a specific set of risk factors rather than a high-dimensional dataset.

#### Evaluation of protein-based prediction models

The performance of each prediction model was evaluated in the testing set by receiver-operating characteristic (ROC) curve analysis, and the DeLong test was used to evaluate differences between AUCs. In addition, we constructed Kaplan-Meier plots to visualize the cumulative incidence of each outcome during follow-up according to proteomic risk score quintiles. We also tested the multivariable-adjusted association of each risk score (as a continuous variable) with their corresponding outcome using multivariable-adjusted Cox regression models adjusted for age, age², self-reported race/ethnicity, the first ten principal components of genetic ancestry, smoking, normalized Townsend deprivation index, BMI, systolic blood pressure, antihypertensive medication use, total cholesterol, HDL cholesterol, cholesterol-lowering medication use, serum creatinine, and prevalent type 2 diabetes. Coronary artery disease, heart failure, atrial fibrillation, and aortic stenosis were included as time-varying covariates. We used the *glmnet*^19^ and *pROC*^20^ packages in R to construct and test all risk scores.

#### Statistical analyses

All tests were two-sided. Data analysis was performed using R (version 4.1.0; *R Project for Statistical Computing*) unless otherwise specified.

### Supplementary Figures

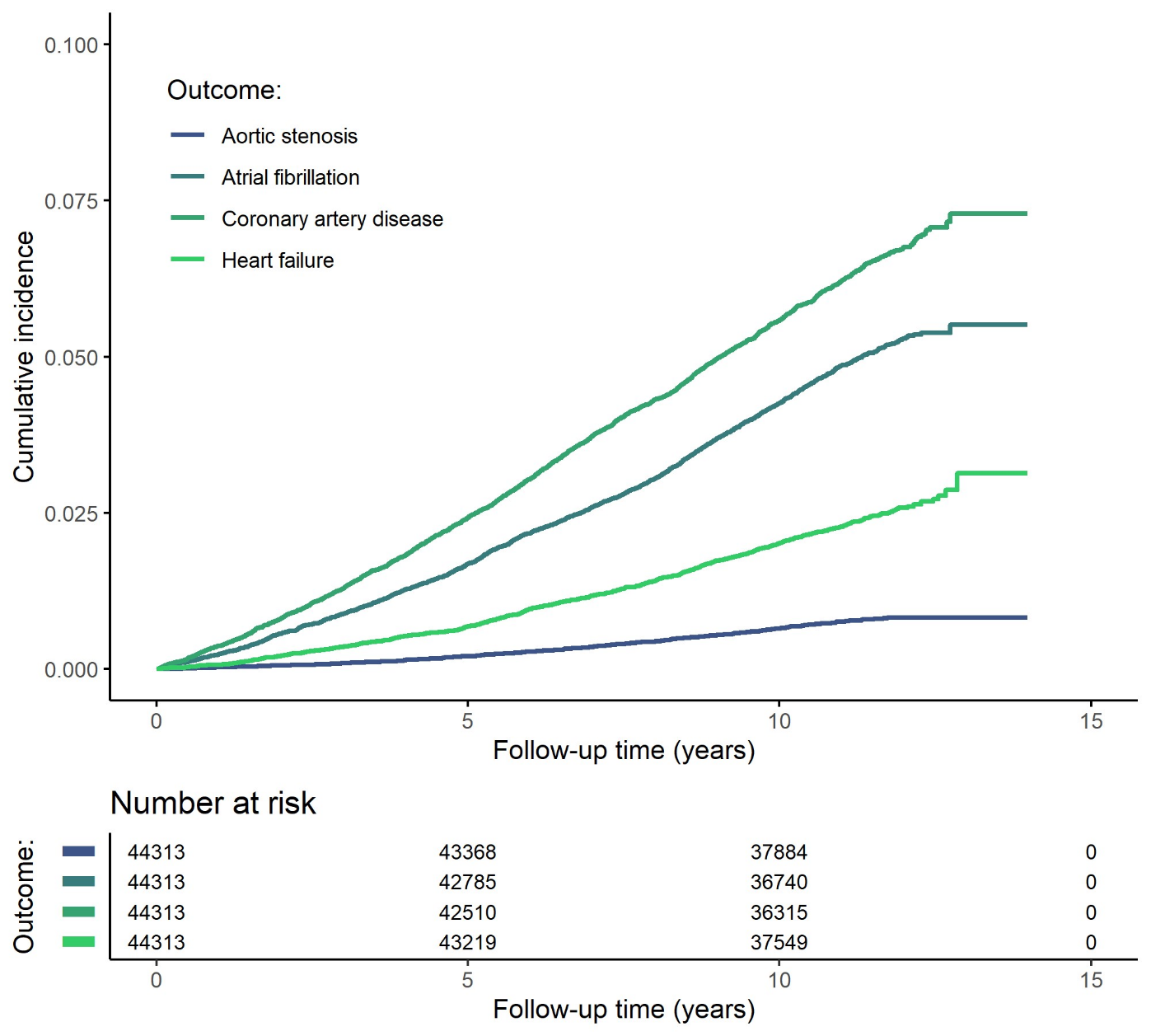

#### Supplementary Fig. 1. Cumulative incidence of coronary artery disease, heart failure, atrial fibrillation, and aotic stenosis during follow-up.

Cumulative incidence plots were constructed using the Kaplan-Meier method. Participants were followed for a median (interquartile range) follow-up of 11.1 (10.4-11.8) years

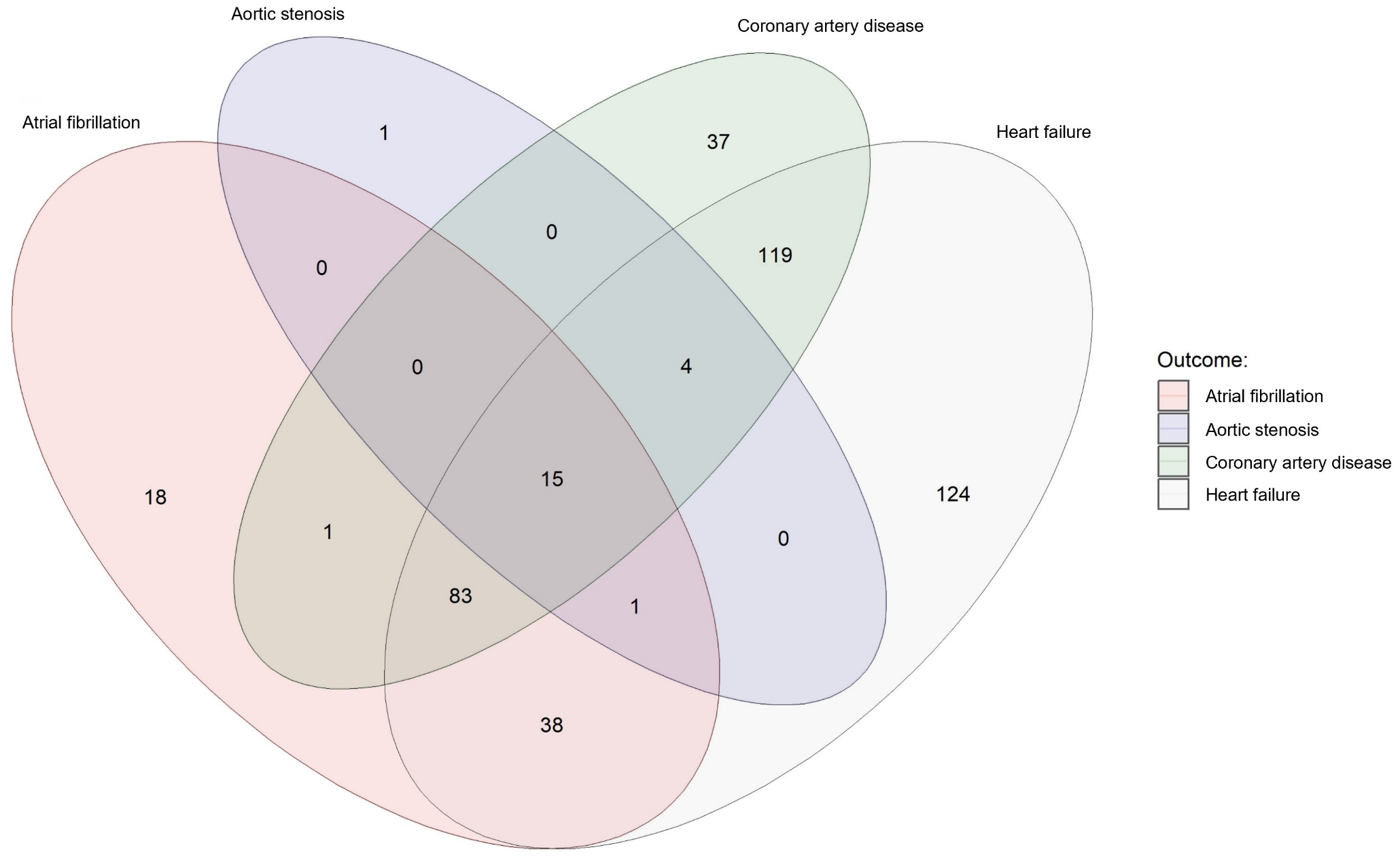

#### Supplementary Fig. 2. Venn diagram showing the number of distinct and shared protein associations across outcomes.

All 441 proteins that were associated with one or more outcomes at Bonferroni-corrected *P*<0.05 are represented in this graph.

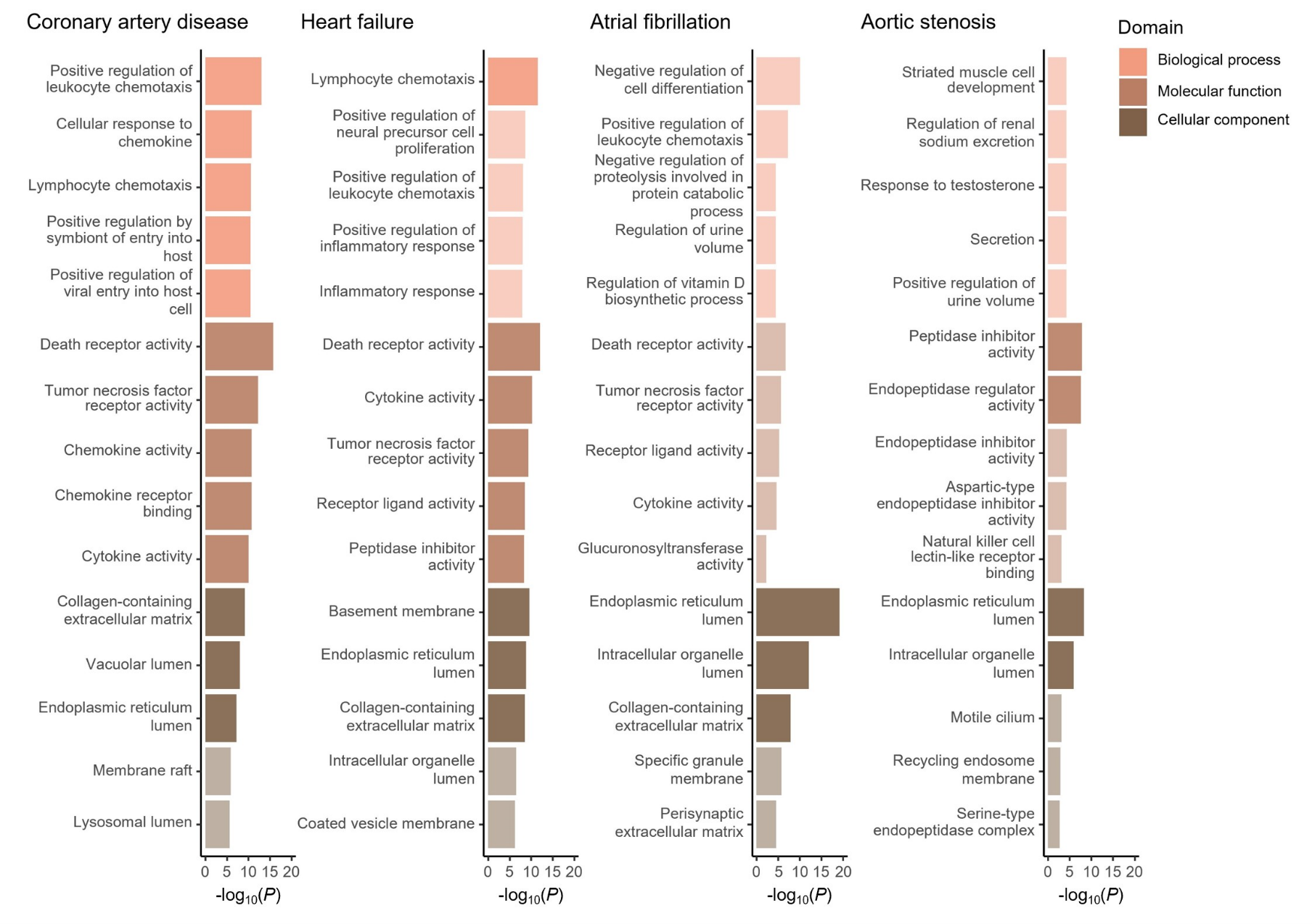

#### Supplementary Fig. 3. Top biological processes, molecular functions, and cellular components enriched among proteins associated with coronary artery disease, heart failure, atrial fibrillation, and aortic stenosis.

Top biological functions, molecular pathways, and cellular components were queried using the *Gene Ontology* resource^21,22^ via *Enrichr.*^23^ Enrichment tests were performed against a background gene set that included the genes corresponding to all 1,459 proteins tested in primary analyses. Gene sets with a false discovery rate-adjusted *P*<0.05 were considered statistically significant. Bright colors indicate statistical significance, whereas dull colors indicate no statistical significance.

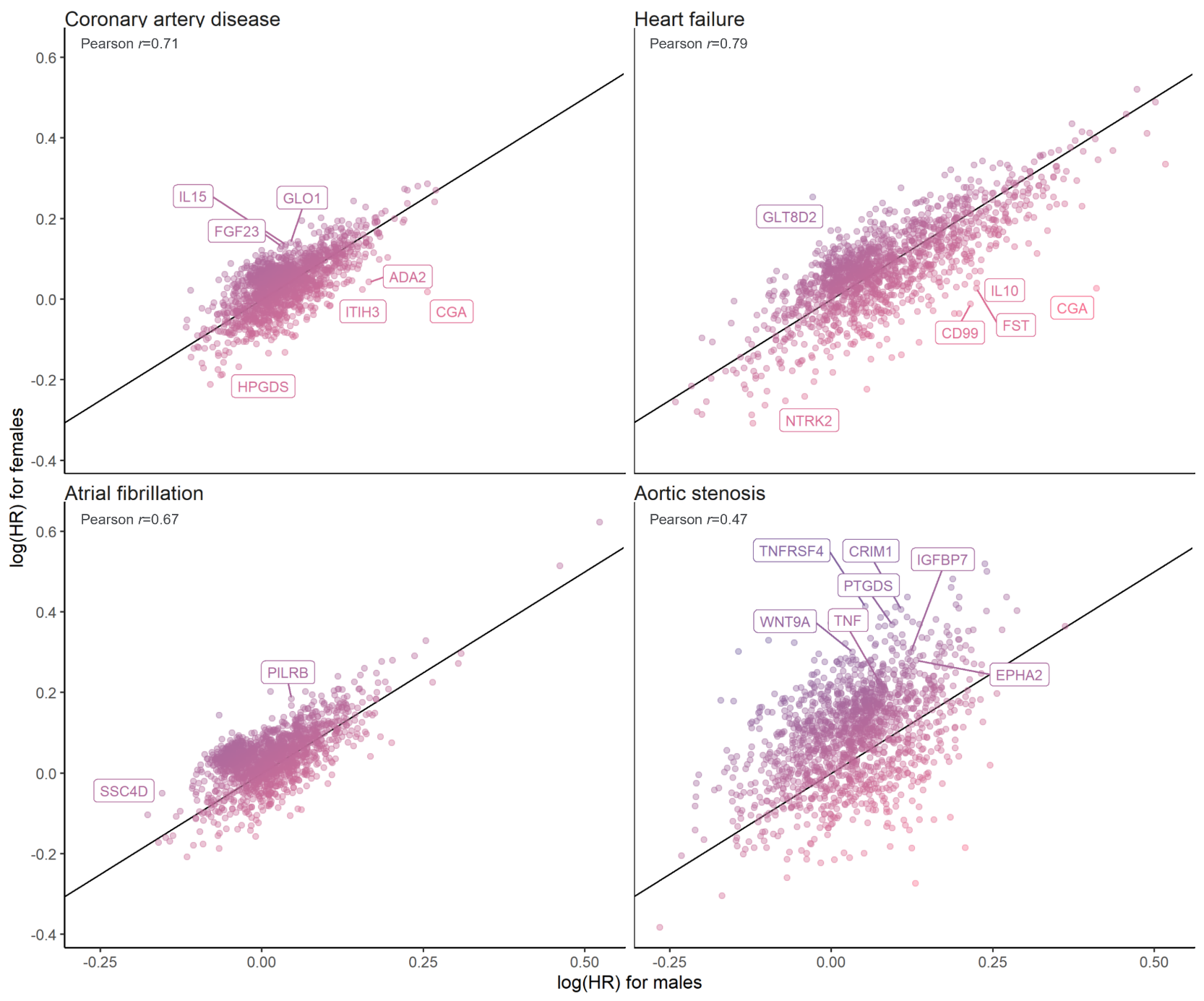

#### Supplementary Fig. 4. Correlation between the effect sizes of protein-disease associations in male vs. female participants.

The scatter plots depict the correlation between the protein-disease associations’ effect sizes (i.e., log[HR]) in female vs. male participants. HR indicates hazard ratio. All estimates were calculated using multivariable-adjusted Cox proportional hazards models, adjusted for age, age², self-reported race/ethnicity, the first ten principal components of genetic ancestry, smoking, normalized Townsend deprivation index, body mass index, systolic blood pressure, antihypertensive medication use, total cholesterol, high-density lipoprotein cholesterol, cholesterol-lowering medication use, serum creatinine, and prevalent type 2 diabetes. In addition, we included the cardiac outcomes that were not tested (e.g., heart failure, atrial fibrillation, and aortic stenosis for incident coronary artery disease models) as time-varying covariates. The labeled protein-disease represent proteins that were associated with the indicated outcome at *P*<0.05/5,836 in one sex without nominal significance (*P*>0.05) in the other sex. In addition, all proteins indicated in color had suggestive evidence for interaction by sex (*P*_interaction_<0.05). HR indicates hazard ratio.

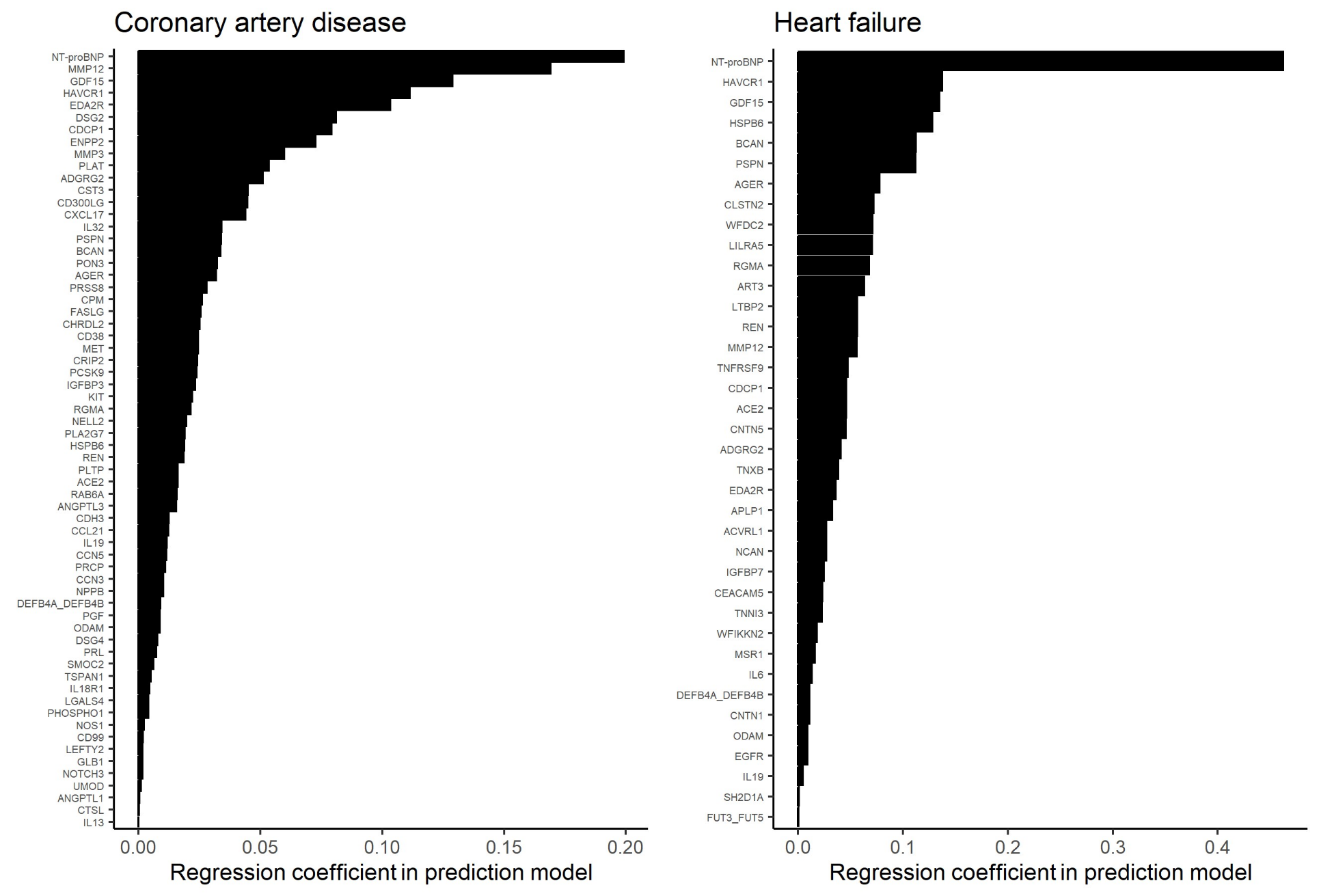

#### Supplementary Fig. 5. Protein weights for the protein-based prediction models of coronary artery disease, heart failure, atrial fibrillation, and aortic stenosis.

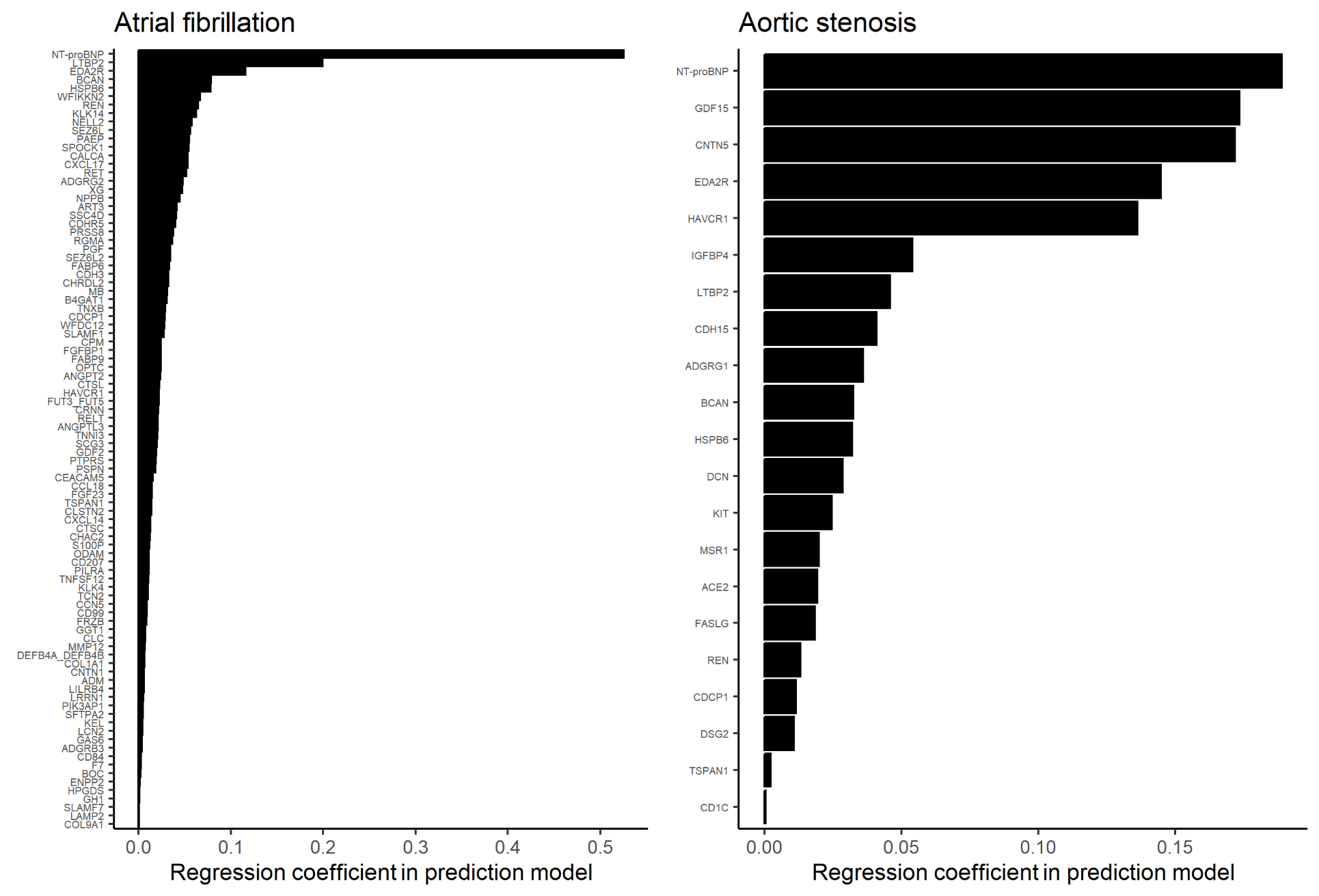

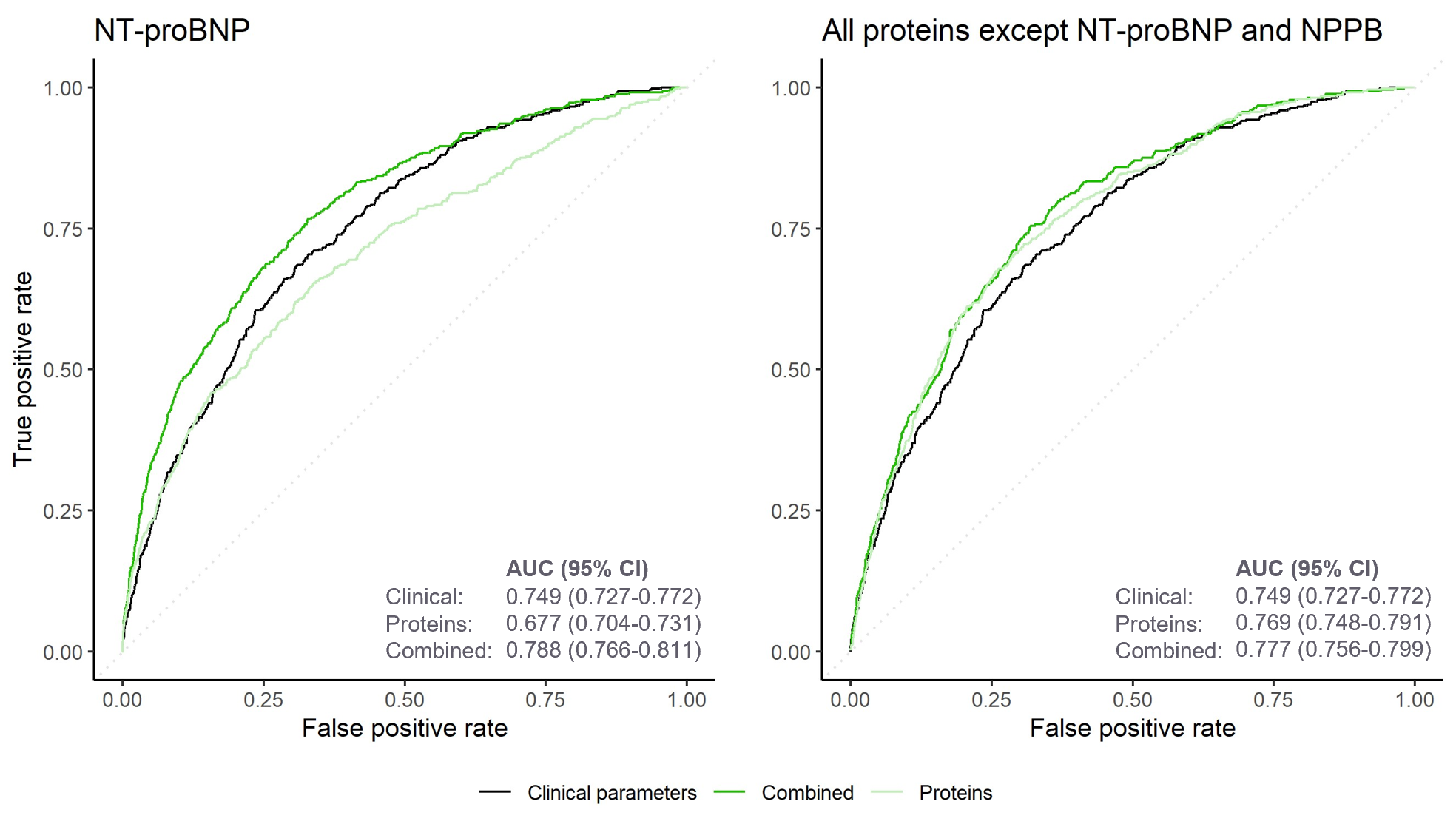

#### Supplementary Fig. 6. Risk prediction of incident atrial fibrillation by risk scores incorporating NT-proBNP and all proteins except NT-proBNP and NPPB.

The receiver-operating characteristics curves depict the accuracy of the clinical, proteomic, and combined risk scores in predicting atrial fibrillation events in the UKB-PPP testing set (*n*=8,863). Areas under the curve (AUCs) and corresponding 95% confidence intervals (95% CIs) quantify the performance of each model. Models with multiple candidate features were constructed using logistic least absolute shrinkage and selection operator (LASSO) models; the combined models included all clinical predictors (see *Methods*) as well as the indicated biomarkers (i.e., NT-proBNP or all proteins except NT-proBNP and NPPB) as potential covariates in the final model. Participants were followed for a median (interquartile range) follow-up of 11.1 (10.4-11.8) years.

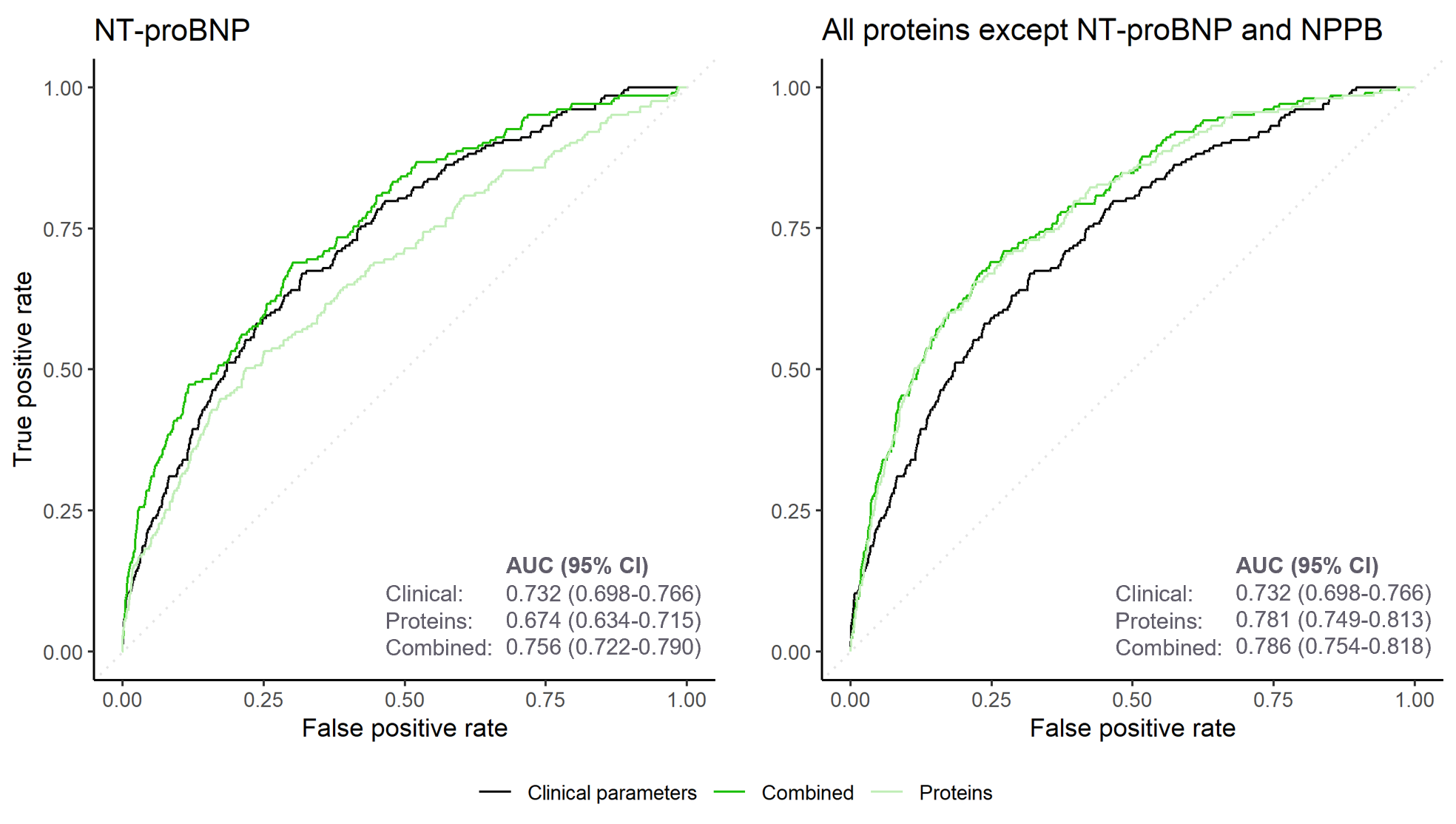

#### Supplementary Fig. 7. Risk prediction of incident heart failure by risk scores incorporating NT-proBNP and all proteins except NT-proBNP and NPPB.

The receiver-operating characteristics curves depict the accuracy of the clinical, proteomic, and combined risk scores in predicting heart failure events in the UKB-PPP testing set (*n*=8,863). Areas under the curve (AUCs) and corresponding 95% confidence intervals (95% CIs) quantify the performance of each model. Models with multiple candidate features were constructed were constructed using logistic least absolute shrinkage and selection operator (LASSO) models; the combined models included all clinical predictors (see *Methods*) as well as the indicated biomarkers (i.e., NT-proBNP or all proteins except NT-proBNP and NPPB) as potential covariates in the final model. Participants were followed for a median (interquartile range) follow-up of 11.1 (10.4-11.8) years.

### Supplementary Tables (legends)

All *Supplementary Tables* can be found in the supplementary Excel file.

#### Supplementary Table 1. Description of the *Olink Explore 1536* proteins included in the present study.

When a single Olink analyte was encoded by two different genes, information for both genes were listed.

#### Supplementary Table 2. Missing protein measurements among the included participants. Missing protein measurements were quantified in UKB-PPP participants who were included in the final study cohort (*N*=44,313).

#### Supplementary Table 3. Associations of circulating protein levels with coronary artery disease, heart failure, atrial fibrillation, and aortic stenosis.

All estimates were calculated using multivariable-adjusted Cox proportional hazards models, adjusted for age, age², sex, self-reported race/ethnicity, the first ten principal components of genetic ancestry, smoking, normalized Townsend deprivation index, body mass index, systolic blood pressure, antihypertensive medication use, total cholesterol, high-density lipoprotein cholesterol, cholesterol-lowering medication use, serum creatinine, and prevalent type 2 diabetes. In addition, we included the cardiac outcomes that were not tested (e.g., heart failure, atrial fibrillation, and aortic stenosis for incident coronary artery disease models) as time-varying covariates. Protein-disease associations with Bonferroni-corrected *P*<0.05 (i.e., *P*<0.05/5,836 or ~8.6×10-6) were considered statistically significant. CI indicates confidence interval; HR, hazard ratio.

#### Supplementary Table 4. Top biological processes, molecular functions, and cellular components enriched among proteins associated with coronary artery disease, heart failure, atrial fibrillation, and aortic stenosis.

Top biological functions, molecular pathways, and cellular components were queried using the *Gene Ontology* resource via *Enrichr*. Enrichment tests were performed against a background gene set that included the genes corresponding to all 1,459 proteins tested in primary analyses. Gene sets with a false discovery rate-adjusted *P*<0.05 were considered statistically significant. The five most strongly enriched pathways per *Gene Ontology* domain are listed for each outcome.

#### Supplementary Table 5. Genetic variants included in the genetic instruments for all proteins with valid *cis*-protein quantitative trait loci (*cis*-pQTLs).

There were a total of 408 proteins with valid *cis*-pQTLs. All positions are expressed using *Genome Reference Consortium Human Build 37* (GRCh37; hg19). pQTL indicates protein quantitative trait locus; SE, standard error.

#### Supplementary Table 6. *F*-statistics and *R*² estimates for the genetic instruments tested in *cis*-Mendelian randomization (MR) analyes.

*F*-statistics were obtained by performing linear regression analyses of a protein’s genetic risk score (as the independent variable) against the measured levels of that protein (as the dependent variable) in the UKB-PPP. Genetic risk scores were calculated as weighted allele scores using the “clumping and thresholding” method, applying the same *P*-value and linkage disequilibrium *R*² thresholds as those used in our primary cis-MR analyses (*P*<5×10^-6^ and *R*²<0.1). All scores were calculated using genotype array data; for proteins where genetic risk score calculation failed, *F*-statistics were estimated using summary statistics as *F*=((*n*-*k*-1)/*k*)×((*R*²)/(1-*R*²)) where *n* indicates the sample size of the original genome-wide association study, *k* the number of variants included in the genetic instrument, and *R*² the variance in the exposure explained by the genetic variants.

#### Supplementary Table 7. Steiger filtering results for protein-disease pairs tested in sensitivity *cis*-Mendelian randomization (MR) analyses.

Steiger filtering was performed and *R*² values were estimated using the *steiger_filtering()* function from the *TwoSampleMR* package in R.

#### Supplementary Table 8. Associations of genetically predicted protein levels with coronary artery disease, heart failure, atrial fibrillation, and aortic stenosis in primary *cis*-Mendelian randomization (MR) analyses.

All analyses represent *cis*-MR analyses performed using the inverse-variance-weighted (IVW) method (for instruments with two or more variants) or the Wald ratio method (for instruments with a single variant). Genetic instruments were constructed using cis-variants associated with circulating protein levels at *P*<1×10^-4^ clumped at *R*²<0.1. All associations are expressed per standard deviation increase in genetically predicted protein levels. CI indicates confidence interval; OR, odds ratio.

#### Supplementary Table 9. Associations of genetically predicted protein levels with coronary artery disease, heart failure, atrial fibrillation, and aortic stenosis in sensitivity Mendelian randomization (MR) analyses using different instrument construction parameters and MR-Egger.

All analyses represent *cis*-MR analyses performed using the indicated methods. Genetic instruments were constructed using *cis*-variants associated with circulating protein levels that were clumped at the indicated *P*-value and *R*² thresholds. Analyses using the inverse-variance-weighted (IVW) method incorporating principal components used genetic instruments that were not clumped against a correlation threshold. Associations were considered robust if the primary analysis was statistically significant (false discovery rate [FDR]-adjusted *P*<0.05) and all sensitivity analyses were directionally consistent. N/A indicates that we were unable to calculate MR estimates (if no variants met the genetic instrument selection criteria). CI indicates confidence interval; LD, linkage disequilibrium; OR, odds ratio.

#### Supplementary Table 10. Associations of genetically predicted protein levels with coronary artery disease, heart failure, atrial fibrillation, and aortic stenosis in one-sample *cis*-Mendelian randomization (MR) analyses.

All proteins with putative causal associations that were robust to the sensitivity analyses presented in **Supplementary Table 9** were tested in one-sample MR analyses. One-sample MR was performed in UKB participants who were not included in the UKB-PPP, were free of cardiac diseases at baseline, and had no missing covariates (*N*=407,230). Associations of the circulating proteins’ genetic scores with cardiac outcomes were tested using Cox regression models adjusted for age, age², sex, race/ethnicity, and the first ten principal components of genetic ancestry. N/A indicates proteins for which genetic score calculation failed. CI indicates confidence interval; HR, hazard ratio; IVW, inverse-variance weighted; LD, linkage disequilibrium; OR, odds ratio.

#### Supplementary Table 11. Evaluation of shared protein quantitative trait loci (pQTLs) between proteins for biomarkers that were robust to sensitivity *cis*-Mendelian randomization (MR) analyses.

Linear regression analyses tested the associations of genetic scores for all proteins surviving upstream sensitivity analyses (**Supplementary Tables 9-10**) with all proteins with putative causal associations in the primary *cis*-MR analyses. Linear regression models were adjusted for age, age², sex, race/ethnicity, and the first ten principal components of genetic ancestry. Associations with Bonferroni-corrected *P*<0.05/13,680 (~3.7×10^-6^) were considered statistically significant. CI indicates confidence interval.

#### Supplementary Table 12. Multivariable-adjusted *cis*-Mendelian randomization (MR) analyses testing the genetic associations of proteins robust to sensitivity analyses with coronary artery disease, heart failure, atrial fibrillation, and aortic stenosis, adjusting for proteins with shared protein quantitative trait loci (pQTLs).

For proteins with genetic instruments that were robust to upstream sensitivity analyses (**Supplementary Tables 9-10**) and significantly associated with one or more other proteins (i.e., “correlated proteins”; **Supplementary Table 11**), we calculated multivariable-adjusted *cis*-MR estimates in a UKB sample excluding UKB-PPP participants, those with cardiac diseases at baseline, and those with missing covariates (*N*=407,230) using Cox regression models. ^a^Cox regression models were adjusted for age, age², sex, race/ethnicity, the first ten principal components of genetic ancestry, and the genetic risk scores of all “correlated” proteins (i.e., the listed proteins). CI indicates confidence interval; HR, hazard ratio.

#### Supplementary Table 13. Colocalization of *cis*-protein quantitative trait loci (pQTLs) with cardiac diseases for protein-diseases associations that were robust to sensitivity Mendelian randomization (MR) analyses.

Colocalization analyses were performed for all protein-disease associations that were robust to sensitivity MR analyses, using *cis*-variants (using a window size of ±200 kilobases) associated with circulating protein levels. The current table shows posterior probabilities for *H_0_* (neither trait has a genetic association in the region), *H_1_* (only trait 1 [i.e., the indicated protein's circulating levels] has a genetic association in the region), *H_2_* (only trait 2 [i.e., the indicated disease] has a genetic association in the region), *H_3_* (both traits are associated but with different causal variants), and *H_4_* (both traits are associated and share a single causal variant).

#### Supplementary Table 14. Sex-stratified associations of circulating protein levels with coronary artery disease, heart failure, atrial fibrillation, and aortic stenosis.

*P*_interaction_ indicates the *P*-value for the interaction term between “sex” and the indicated protein on the corresponding outcome. All associations were tested using multivariable-adjusted Cox proportional hazards models, adjusted for age, age², self-reported race/ethnicity, the first ten principal components of genetic ancestry, smoking, normalized Townsend deprivation index, body mass index, systolic blood pressure, antihypertensive medication use, total cholesterol, high-density lipoprotein cholesterol, cholesterol-lowering medication use, serum creatinine, and prevalent type 2 diabetes. In addition, we included the cardiac outcomes that were not tested (e.g., heart failure, atrial fibrillation, and aortic stenosis for incident coronary artery disease models) as time-varying covariates. Interaction models were additionally adjusted for sex and circulating levels of the indicated protein.

#### Supplementary Table 15. Coefficients for the covariates included in the protein-based, clinical, and combined prediction models for coronary artery disease, heart failure, atrial fibrillation, and aortic stenosis.

All risk scores were constructed in the training set using logistic least absolute shrinkage and selection operator (LASSO) regression analyses with 10-fold cross-validation in the UKB-PPP training set (80%: n=35,450). N/A indicates not applicable; SD, standard deviation.

#### Supplementary Table 16. Diagnostic and procedural codes used to define disease-related covariates and outcomes.

Prevalent disease status was ascertained by self-report at enrolment or qualifying diagnostic or procedural codes. Incident disease status was ascertained by qualifying diagnostic or procedural codes. ICD indicates *International Classification of Diseases*; OPCS indicates *Office of Population Censuses and Surveys Classification of Surgical Operations and Procedures*.
